# Modelling the relative influence of socio-demographic variables on post-acute COVID-19 quality of life

**DOI:** 10.1101/2024.02.21.24303099

**Authors:** Tigist F. Menkir, Barbara Wanjiru Citarella, Louise Sigfrid, Yash Doshi, Luis Felipe Reyes, Jose A. Calvache, Anders Benjamin Kildal, Anders B. Nygaard, Jan Cato Holter, Prasan Kumar Panda, Waasila Jassat, Laura Merson, Christl A. Donnelly, Mauricio Santillana, Caroline Buckee, Stéphane Verguet, Nima S. Hejazi, The ISARIC Clinical Characterisation Group

## Abstract

**Introduction:** Long-term COVID-19 complications are a globally pervasive threat, but their social determinants are often understudied relative to clinical risk factors. Thus, the role that clinical comorbidities play in determining disparate long COVID outcomes across socio-demographic groups is relatively unknown. Here, we ranked social and clinical predictors of quality of life (QoL) with long COVID and measured the extent to which clinical intermediates explain any observed relationships between social factors and long COVID QoL.

**Methods:** We used demographic, comorbidity, treatment, and quality of life data from acute case reporting forms and follow-up surveys collected for a multinational prospective cohort study, with focus on data from Norway (n=1,672), the UK (n=1,064), and Russia (n=1,155). We considered the social factors employment status, educational attainment, and female sex and defined our primary outcome as QoL utility scores among subjects reporting any long COVID-associated symptoms.

**Results:** We found that, in addition to age, neuropsychological, and rheumatological comorbidities, educational attainment, employment status, and sex were consistently identified as top predictors of long COVID-associated QoL utility scores. Furthermore, 98.2% (95% CI: 87.3%, 100%) and 89.4% (74.6%, 99.6%) of the positive adjusted associations between high educational attainment or full-time employment and long COVID QoL utility scores remained unexplained by key long COVID-predicting comorbidities in Norway and the UK. The same was true for 84.2% (46%, 100%), 67.6% (45.5%, 89.7%), and 87.2% (71.3%, 100%) of the negative adjusted associations between female sex and long COVID QoL utility scores in Norway, the UK, and Russia.

**Conclusion:** Socio-economic proxies and sex are strong predictors of long COVID QoL independent of commonly emphasized comorbidity pathways and warrant increased attention in interventions focused on mitigating long COVID burden. Long COVID management efforts that target social determinants should be tailored to a specific country context, given the heterogeneity in findings observed across settings.

**What is already known on this topic:**

- Clinical comorbidities like asthma, chronic cardiac disease, and diabetes, are well-established long COVID risk factors; social factors, namely socioeconomic
- disadvantage and gender, have also been identified as important sources of long COVID risk; the relative rankings of these factors in differentiating quality of life with long COVID is less recognized
- Furthermore, key clinical predisposing factors have been shown to play a minimal role in confounding observed social disparities in long COVID risk; their potential role as explanatory intermediates is generally underexplored

**What this study adds:**

- Evaluates both social and clinical factors (individually and grouped by shared mechanisms) as predictors of long COVID-associated quality of life, an outcome measure that can capture a gradient in experiences with the condition
- Importantly, subverts the commonplace emphasis on differences in comorbidity burden explaining observed disparities in health outcomes by quantifying the proportion of relationships between different social factors and long COVID quality of life that are *unexplained* by clinical intermediates, applying flexible statistical mediation approaches

**How this study might affect research, practice or policy:**

- Motivates broadening the focus of long COVID prevention efforts to look beyond simply targeting clinical intermediates as a way to fully resolve social disparities and instead also explore the role of other upstream societal sources of disadvantage, for example, access to public health resources and clinical services or discrimintion

## Introduction

Long-term COVID-19 sequelae, referred to as long COVID, have contributed to a growing public health crisis since 2020. Long COVID encompasses unexplainable symptoms which persist at least three months after an infection, occurring over two or more months (1). Its widespread presence and impacts have been immense: a multinational study found that nearly half of individuals who were previously infected with SARS-CoV-2 went on to experience long-term symptoms four months after infection (2); further, an estimated 59% of previously infected participants reported a reduced quality of life (QoL) (3,4). Prior work has focused on identifying a myriad of clinical risk factors for post-COVID-19 conditions, including co-infections and pre-existing conditions, vaccination status, age, and female sex (4–7). Key correlates of long COVID sequelae consistently identified include obesity, asthma and other pulmonary diseases, chronic cardiac disease, diabetes, and smoking (6,7).

Beyond clinical factors, social vulnerabilities are often critical determinants of differential disease burden overall (8–11). Such inequities are attributed to broader challenges in access to health services and an array of health-threatening exposures, including but not limited to food and housing insecurity, financial discrimination, and air pollution (8–11).

While there have been efforts to examine social factors potentially linked to long-term symptoms of COVID-19 (7,12–20), findings on these relationships have been somewhat mixed (13, 18). For instance, a 2021 study in the United Kingdom (UK) found that living in high-deprivation settings was associated with both a higher and lower odds of symptom persistence, depending on the measure of deprivation index used (18), and a 2021 study in Michigan (USA) found that lower income was both significantly and non-significantly correlated with long COVID symptoms’ prevalence, depending on the post-illness duration considered (13). It is also important to highlight that many of these studies rely on self-reported binary measures of post-COVID recovery, which do not capture nuanced experiences in recovery.

Given this context, we aimed to complement efforts centered on uncovering disparities in long COVID outcomes by leveraging a large, multi-national prospective cohort study. Specifically, we formally assessed how a diverse group of exposures may differentially predict long COVID QoL, reasoning that factors like socio-economic status (SES) would be as much or more critical predictors than important comorbidities, as has been recently illustrated for related outcomes, such as “healthy aging” (21). To further isolate the unique contribution of social determinants of health, we then evaluated the degree by which clinical intermediates may explain social disparities in long COVID QoL, hypothesising that differences in comorbidity risk may not fully account for these disparities. To do so, we apply a similar mediation-centred lens to that of Vahidy et al. (17) and Lu et al. (22). However, rather than independently evaluating the role of various clinical mediators (17) or social factors as mediators (22), we measured how much social variables’ associations with long COVID QoL *cannot* be explained by a collective set of comorbidities.

## Methods

This study uses data from the International Severe Acute Respiratory and emerging Infection Consortium’s (ISARIC) multi-cohort consortium (23). This prospective study across 76 countries collected demographic and medical data during acute SARS-CoV-2 infection from 2020 to 2022, based on hospital admissions case reporting forms, with a subset of sites assessing participants through detailed surveys starting from one-to-three months post-infection and periodically thereafter (23,24). Complete details on the study design and recruitment procedures can be found in the published follow-up protocol (23,24). Within this multi-national cohort, we subset to countries with data available on SES, age, sex, quality of life, and comorbidities, with combined demographic, comorbidity, and QoL datasets yielding sample sizes of at least n=1000. The countries meeting this criterion were Norway (n=1,672), the UK (n=1,064), and Russia (n=1,155). Our study population consisted of subjects self-reporting one or more long COVID-associated symptoms not present prior to illness, at least one month following infection. Health utility values were obtained from responses to the EQ-5D-5L survey in follow-up forms, eliciting self-reported rankings of problem severity across five health dimensions (mobility, self-care, usual activities, pain/discomfort, and anxiety/depression) (25), relative to both pre-COVID and current experiences.

We incorporated covariate data on age, female sex-at-birth, SES proxies, clinical comorbidities, treatments, and COVID-19 severity. SES indicators were selected based on data availability within each cohort: educational attainment in years for Norway, and employment status categories for the UK and Russia. For our mediation analyses, SES indicators were dichotomized as follows: high versus low educational attainment (quintiles 3–5 versus quintiles 1–2) in Norway, and full-time versus other employment status in the UK and Russia. Mediators of interest were selected based on a literature review of studies identifying key long COVID predictors (6,7), in accordance with the data available for each country. A full list of mediators for each cohort is provided in the Supplementary Appendix.

We estimated QoL scores among those classified as experiencing long COVID, as defined above. Composite QoL utility scores were computed following standard practice, incorporating relevant country-specific external weights for each EQ-5D-5L response. Utility scores range from −1 to 1, where values approaching 1 indicate fewer health problems and greater QoL, values approaching 0 indicate poorer QoL, and negative values reflect a health state considered “worse than death” (26). Given the absence of subject-specific data on symptom duration, we do not present time-adjusted QoL measures, as doing so would require the simplifying assumption of equivalent symptom duration across participants. We similarly did not compare pre-versus post-COVID changes in QoL, as retrospective recall of QoL experiences several months prior is likely to introduce bias.

For our first aim, we identified social predictors of long COVID QoL utility scores using random forest ensemble learners (27), fit separately for each country using all available clinical and demographic data. We implemented three distinct approaches to capture a range of assumptions about variable relationships: variables were treated individually (RF #1), pre-grouped based on subject matter knowledge (RF #2), or grouped algorithmically via hierarchical clustering (RF #3) (28). Variable importance was quantified as the percent increase in mean squared error (MSE) upon variable permutation for RF #1 and #2, and as the frequency of variable selection for RF #3.

For our second aim, we applied statistical techniques developed for causal mediation analysis to examine how clinical intermediates explain observed relationships between our social factors (SES indicators and female sex) with long COVID QoL utility scores. Specifically, for each cohort, we quantified how much the association between a given social factor and long COVID QoL utility scores, adjusted for key demographic variables, operates through the clinical intermediates (the natural indirect effect or NIE) versus through other pathways (the natural direct effect or NDE). Consequently, we were able to quantify what proportion of the total observed associations between the social factors and long COVID QoL were explained by, and unexplained by, the clinical intermediates examined.

To estimate these quantities, we used a flexible mediation analysis approach that incorporates a variety of regression-based and nonparametric machine learning algorithms to provide robust estimates that do not rely on strict modeling assumptions about the nature of the relationship between the social exposures, clinical intermediates, and long COVID QoL (29–32). Extensive methodological details, including the assumptions underlying a causal interpretation of our results, are provided in the Supplementary Appendix. All results are reported with 95% confidence intervals.

## Data availability statement

The data that underpin this analysis are highly detailed clinical data on individuals hospitalised with COVID-19. Due to the sensitive nature of these data and the associated privacy concerns, they are available via a governed data access mechanism following review of a data access committee. Data can be requested via the IDDO COVID-19 Data Sharing Platform (http://www.iddo.org/covid-19). The Data Access Application, Terms of Access and details of the Data Access Committee are available on the website. Briefly, the requirements for access are a request from a qualified researcher working with a legal entity who have a health and/or research remit; a scientifically valid reason for data access which adheres to appropriate ethical principles.The full terms are at: https://www.iddo.org/document/covid-19-data-access-guidelines.

## Code availability statement

All code (with the exception of code used to process the individual datasets) is available at: https://github.com/goshgondar2018/social_long_covid.

## Ethics statement

The ISARIC-WHO Clinical Characterisation Protocol was approved by the World Health Organization Ethics Review Committee (Ref RPC571/RPC572 25APR13). Institutional Ethics Committee approval was additionally obtained by participating sites including the South Central Oxford C Research Ethics Committee in England (Ref 13/SC/0149), Leeds West Research Ethics Committee (Ref 20/YH/0225), and the Scotland A Research Ethics Committee (Ref 20/SS/0028) for the United Kingdom and the Human Research Ethics Committee (Medical) of the University of the Witwatersrand (Ref M2010108) for South Africa, representing the majority of the data. Other institutional and national approvals were obtained by participating sites as per local requirements.

## Patient & public involvement statement

Patients and the public were therefore not involved in the design of the acute phase rapid response research. However, patients and people living with long Covid were involved in the design, conduct and interpretation of the follow up study. The CRF was piloted with patients presenting in clinics in three countries and the feedback incorporated into the final version. This included suggestions on the data on symptoms collected, how questions were asked and the patient information.

## Results

*Throughout this section, we borrow the terms “direct effect” and “indirect effect” from the causal mediation analysis literature, using these as shorthand for decompositions of adjusted associations operating outside of and through the clinical intermediates, respectively; a causal interpretation of these quantities requires additional untestable identification assumptions, which we do not make but review for completeness in the Supplementary Appendix*.

### Norway

The median of long COVID QoL utility scores was 0.944 (interquartile range (IQR): 0.858, 0.986). There was no broadly consistent trend in QoL utility scores across quintiles of educational attainment, although the lowest scores occurred in the bottom two quintiles. The greatest differences in scores occurred between quintiles 4 and 2 and between 4 and 1. Estimated scores among males slightly exceeded that of females (p<0.001). This cohort was the youngest, with a mean age of 51.8 years (SD: 13.6 years). The most commonly reported comorbidity was asthma (22%).

Among the leading individual predictors of long COVID QoL utility scores, anxiety/depression ranked first, followed by educational attainment, rheumatological disorder, and age (Figure 1a). When considering pre-grouped variables as predictors, groups containing all socio-demographic variables, i.e., educational attainment indicators and sex, ranked below groups containing psychological disorder and chronic neurologic disorder (Figure 1b). The alternative grouped approach we applied largely corroborated these orderings, where psychological disorder, rheumatological disorder, chronic neurological disorder, and asthma were the most consistently selected variables within grouped variables identified as important, followed by educational attainment (in years) and an indicator for the fifth quintile of educational attainment (Figure 1c).

**Table 1:** Summary of demographic variables (excluding SES proxies) and common comorbidities in the final study populations for each cohort, post-missing data imputation.

|  | Norway<br>(N=1672) | Russia<br>(N=1155) | UK<br>(N=1064) | Overall<br>(N=3891) |
| --- | --- | --- | --- | --- |
| **AGE** |  |  |  |  |
| Mean (SD) | 51.8 (13.6) | 59.6 (14.4) | 59.0 (12.6) | 56.1 (14.1) |
| Median [Min, Max] | 52.0 [17.0, 96.0] | 59.0 [20.0, 100] | 59.0 [19.0, 91.0] | 56.0 [17.0, 100] |
| **SEX** |  |  |  |  |
| Female | 1138 (68.1%) | 688 (59.6%) | 434 (40.8%) | 2260 (58.1%) |
| Male | 534 (31.9%) | 467 (40.4%) | 630 (59.2%) | 1631 (41.9%) |
| **ASTHMA** |  |  |  |  |
| ABSENT | 1305 (78.1%) | 1076 (93.2%) | 818 (76.9%) | 3199 (82.2%) |
| PRESENT | 367 (21.9%) | 79 (6.8%) | 246 (23.1%) | 692 (17.8%) |
| **CHRONIC CARDIAC DISEASE (NOT HYPERTENSION)** |  |  |  |  |
| ABSENT | 1547 (92.5%) | 877 (75.9%) | 952 (89.5%) | 3376 (86.8%) |
| PRESENT | 125 (7.5%) | 278 (24.1%) | 112 (10.5%) | 515 (13.2%) |
| **CHRONIC HAEMATOLOGICAL DISEASE** |  |  |  |  |
| ABSENT | 1642 (98.2%) | 1137 (98.4%) | 1032 (97.0%) | 3811 (97.9%) |
| PRESENT | 30 (1.8%) | 18 (1.6%) | 32 (3.0%) | 80 (2.1%) |
| **CHRONIC KIDNEY DISEASE** |  |  |  |  |
| ABSENT | 1635 (97.8%) | 1081 (93.6%) | 1004 (94.4%) | 3720 (95.6%) |
| PRESENT | 37 (2.2%) | 74 (6.4%) | 60 (5.6%) | 171 (4.4%) |
| **CHRONIC NEUROLOGICAL DISORDER** |  |  |  |  |
| ABSENT | 1556 (93.1%) | 1087 (94.1%) | 1013 (95.2%) | 3656 (94.0%) |
| PRESENT | 116 (6.9%) | 68 (5.9%) | 51 (4.8%) | 235 (6.0%) |
| **CHRONIC PULMONARY DISEASE (NOT ASTHMA)** |  |  |  |  |
| ABSENT | 1602 (95.8%) | 1037 (89.8%) | 875 (82.2%) | 3514 (90.3%) |
| PRESENT | 70 (4.2%) | 118 (10.2%) | 189 (17.8%) | 377 (9.7%) |
| **DIABETES MELLITUS (TYPE 2)** |  |  |  |  |
| ABSENT | 1542 (92.2%) | 942 (81.6%) | 846 (79.5%) | 3330 (85.6%) |
| PRESENT | 130 (7.8%) | 213 (18.4%) | 218 (20.5%) | 561 (14.4%) |
| **DIABETES MELLITUS (TYPE NOT SPECIFIED)** |  |  |  |  |
| ABSENT | 1646 (98.4%) | 934 (80.9%) | 828 (77.8%) | 3408 (87.6%) |
| PRESENT | 26 (1.6%) | 221 (19.1%) | 236 (22.2%) | 483 (12.4%) |
| **HYPERTENSION** |  |  |  |  |
| ABSENT | 1322 (79.1%) | 475 (41.1%) | 685 (64.4%) | 2482 (63.8%) |
| PRESENT | 350 (20.9%) | 680 (58.9%) | 379 (35.6%) | 1409 (36.2%) |
| **MALIGNANT NEOPLASM** |  |  |  |  |
| ABSENT | 1626 (97.2%) | 1099 (95.2%) | 1034 (97.2%) | 3759 (96.6%) |
| PRESENT | 46 (2.8%) | 56 (4.8%) | 30 (2.8%) | 132 (3.4%) |
| **OBESITY** |  |  |  |  |
| ABSENT | 1638 (98.0%) | 878 (76.0%) | 1006 (94.5%) | 3522 (90.5%) |
| PRESENT | 34 (2.0%) | 277 (24.0%) | 58 (5.5%) | 369 (9.5%) |
| **RHEUMATOLOGICAL DISORDER** |  |  |  |  |
| ABSENT | 1356 (81.1%) | 1090 (94.4%) | 884 (83.1%) | 3330 (85.6%) |
| PRESENT | 316 (18.9%) | 65 (5.6%) | 180 (16.9%) | 561 (14.4%) |
| **SMOKING** |  |  |  |  |
| ABSENT | 1627 (97.3%) | 1011 (87.5%) | 958 (90.0%) | 3596 (92.4%) |
| PRESENT | 45 (2.7%) | 144 (12.5%) | 106 (10.0%) | 295 (7.6%) |
| **Long COVID utility scores** |  |  |  |  |
| Mean (SD) | 0.882 (0.162) | 0.926 (0.134) | 0.738 (0.258) | 0.855 (0.201) |
| Median [Min, Max] | 0.944 [-0.167, 1.00] | 0.966 [-0.590, 1.00] | 0.809 [-0.179, 1.00] | 0.930 [-0.590, 1.00] |

**Figure 1a.**
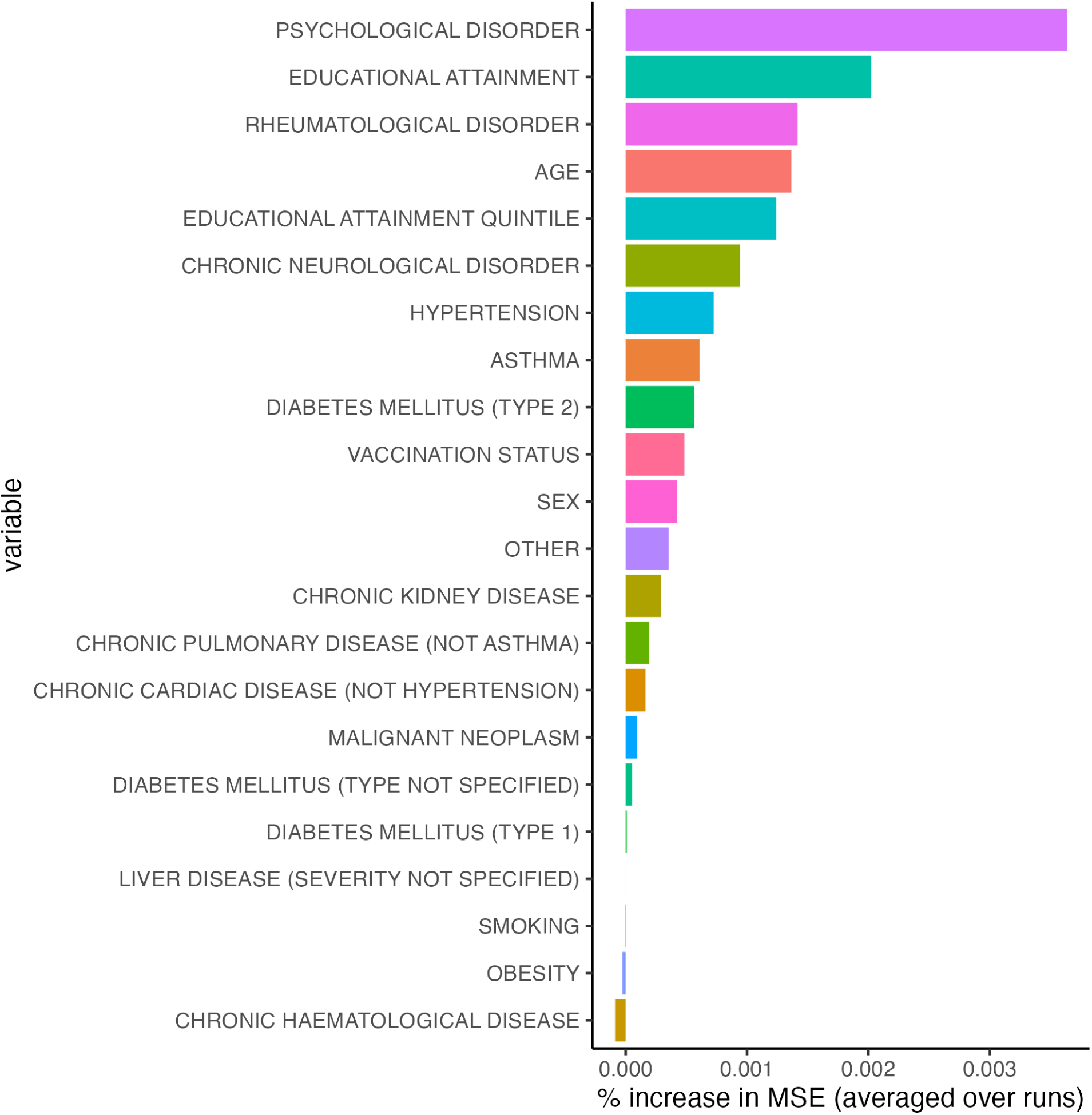
Estimated variable importance measures, i.e. % increase in mean squared error or MSE, from individual random forest implementation (RF #1) for Norway. Variables with negative % MSE values are considered unimportant.

**Figure 1b.**
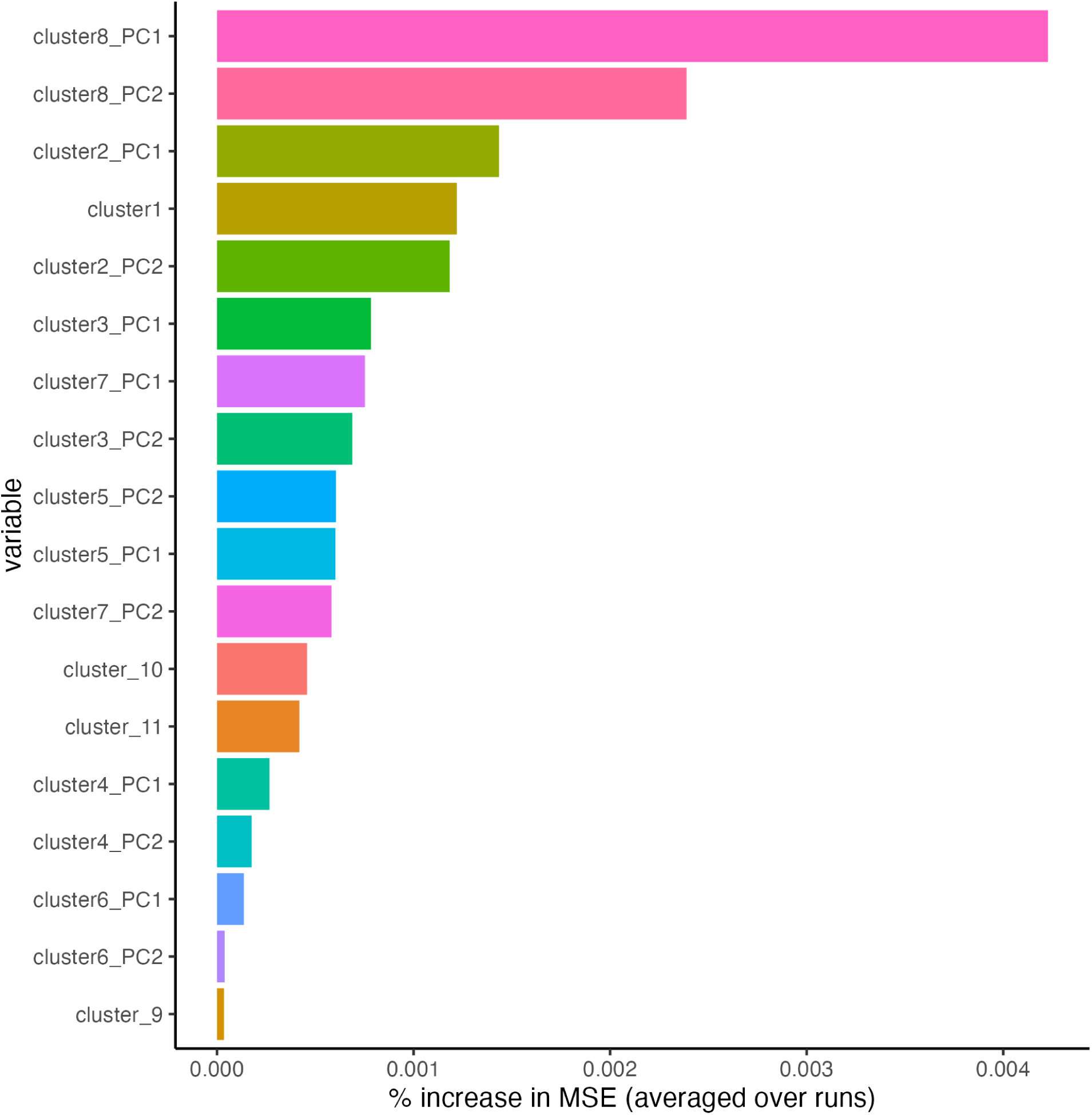
Estimated variable importance measures, i.e. % increase in mean squared error or MSE, from pre-grouped random forest implementation (RF #2) for Norway. Rows indicate cluster names (a full list of variables belonging to each cluster can be found in Supplementary Table S3) and corresponding principal components summarizing the information contained in the cluster, if the cluster consists of multiple variables. PC1 denotes principal component 1 and PC2 denotes principal component 2.

**Figure 1c.**
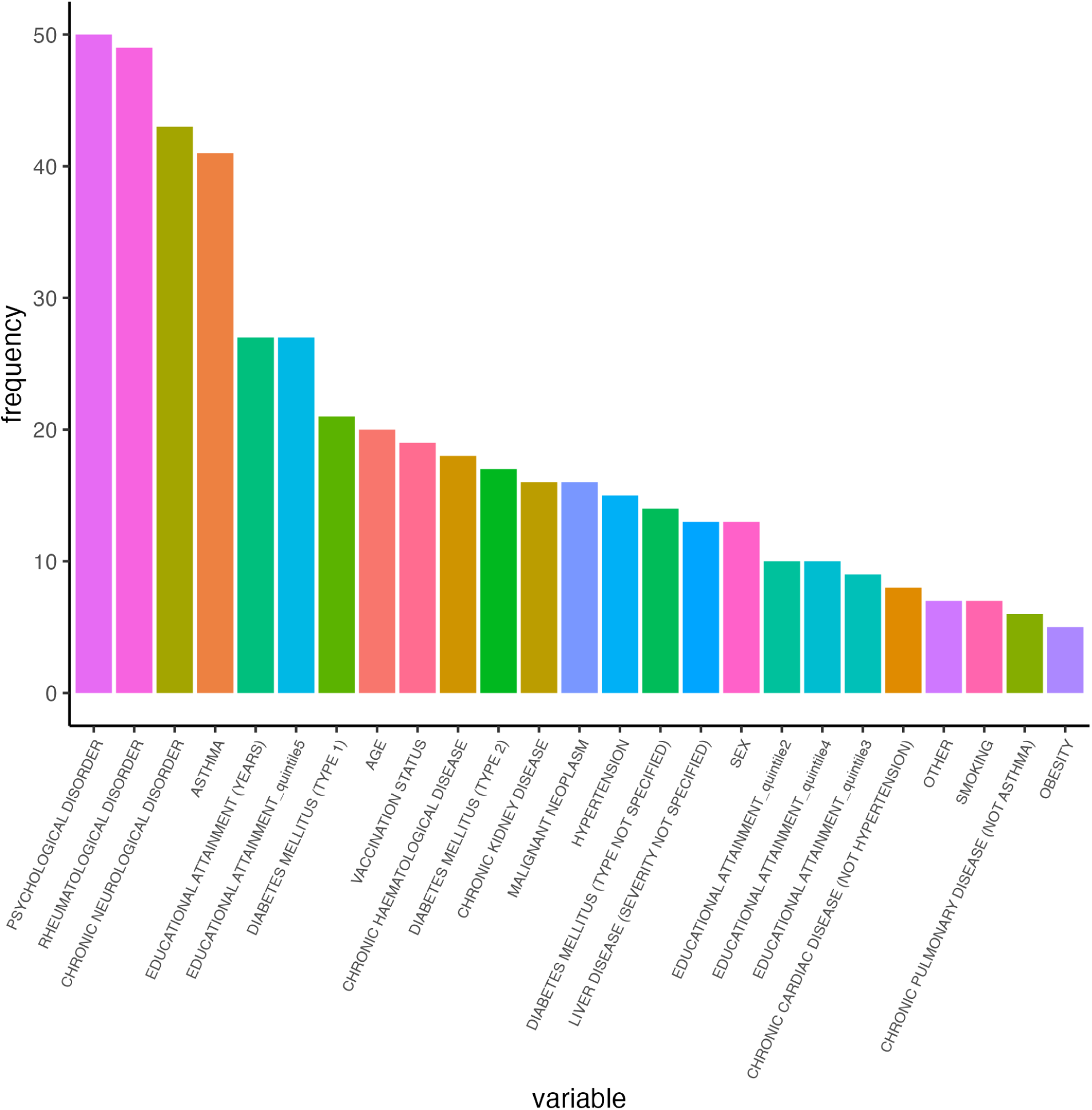
Number of times (frequency) each variable appears in clusters selected as most informative for each run of the model-grouped procedure (RF #3) for Norway.

Higher educational attainment was positively associated with long COVID QoL. We estimated that falling in the top two quintiles of educational attainment was significantly associated with 0.0324 (0.0186, 0.0462) additional units of utility, on average, via the direct effect, and 0.000722 (-0.00333, 0.00478) additional units of utility (non-significant), via the indirect effect, corresponding to a proportion non-mediated of 0.982 (0.873, 1). That is, 98.2% of the positive adjusted association between high educational attainment and long COVID QoL utility scores could not be explained by the included intermediates and is thus attributed to other mechanisms. We obtained consistent directionality in findings for pairwise comparisons of quintiles 4 and 1 (p=0.042), quintiles 4 and 2 (p<0.001), quintiles 5 and 1 (p=0.33), and quintiles 5 and 2 (p=0.0067).

Female sex and QoL utility scores were significantly negatively associated, with an estimated direct effect of -0.0181 (-0.0326, -0.00365), indirect effect of -0.00411 (-0.0115, 0.0033) and proportion non-mediated of 0.842 (0.463,1). In other words, 84.2% of the negative association between female sex and long COVID QoL was unaccounted for by the clinical intermediates examined.

### UK

The median (IQR) of long COVID QoL utility scores was 0.809 (0.637, 0.937). Employment status was skewed towards full-employment (51%), retirement (30%), part-time employment (10%) and unemployment (7%). Estimated QoL utility scores were greatest among participants who reported being furloughed, students, or full-time employees and lowest among those in the unemployed and retired categories. Scores were also slightly higher among males (p<0.001). This cohort was skewed towards older adults (mean (SD): 59.0 (12.6) years) and the most commonly reported comorbidity was hypertension (36%).

Employment status was the leading predictor for long COVID QoL utility scores, followed by anxiety/depression, age, chronic neurological disorder, and rheumatological disorder (Figure 2a). Sex followed in the rankings, which, along with the acute COVID-19 severity indicator, fell among the top ten predictors (Figure 2a). The grouped approach further supported the predictive role of employment status and sex as a group, with groups containing the socio-demographic variables leading the rankings, closely followed by the group containing mental health and neurological disorders (Figure 2b). Age alone was also an influential predictor, even in comparison to grouped factors (Figure 2b). Findings from the model-grouped approach aligned well with these results, with psychological disorder, chronic neurological disorder, employment status indicators being the most commonly selected variables, followed by age, severity indicator and rheumatological disorder, across key groups (Figure 2c).

**Figure 2a.**
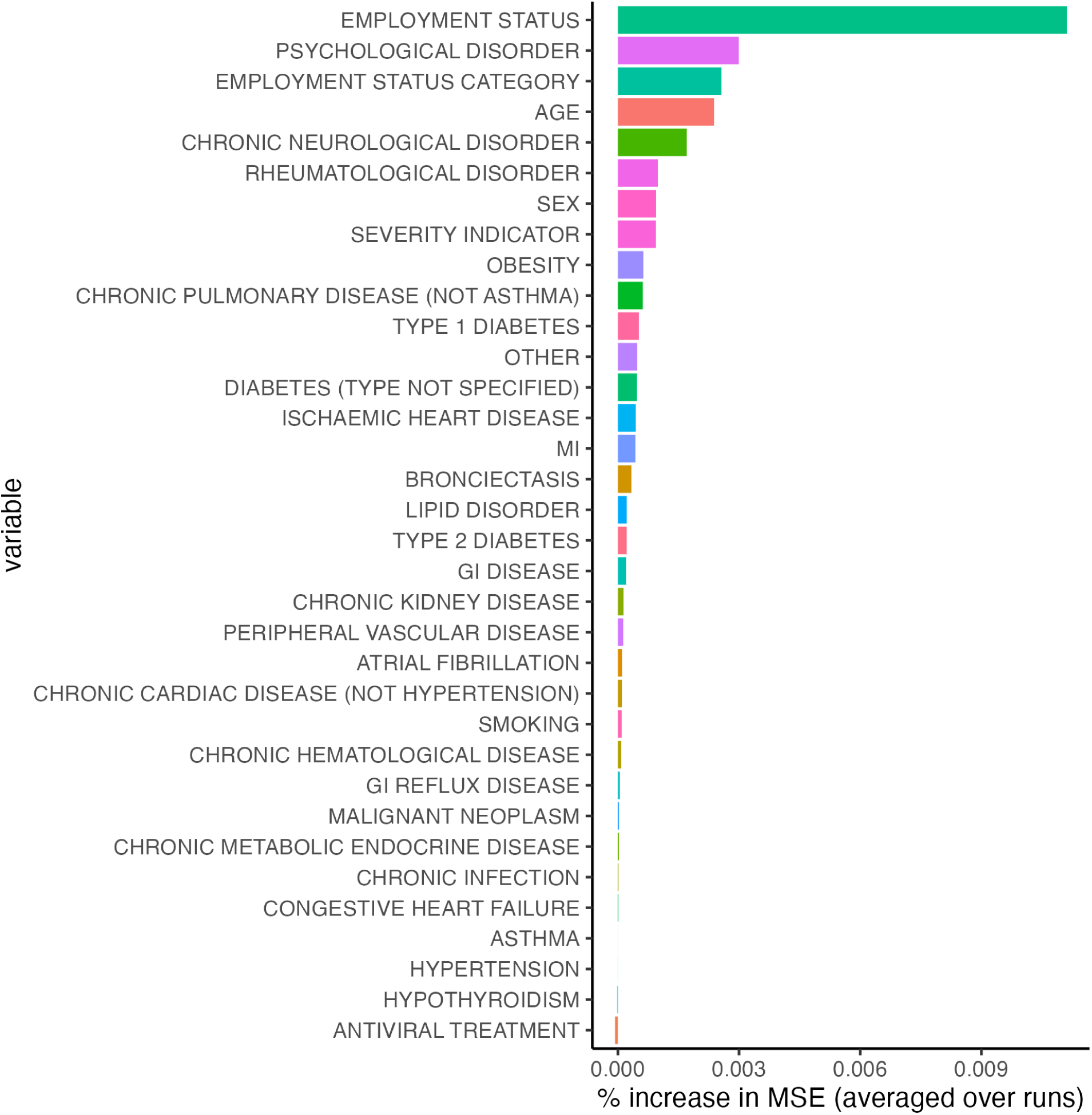
Estimated variable importance measures, i.e. % increase in mean squared error or MSE, from individual random forest implementation (RF #1) for the UK. Variables with negative % MSE values are considered unimportant.

**Figure 2b.**
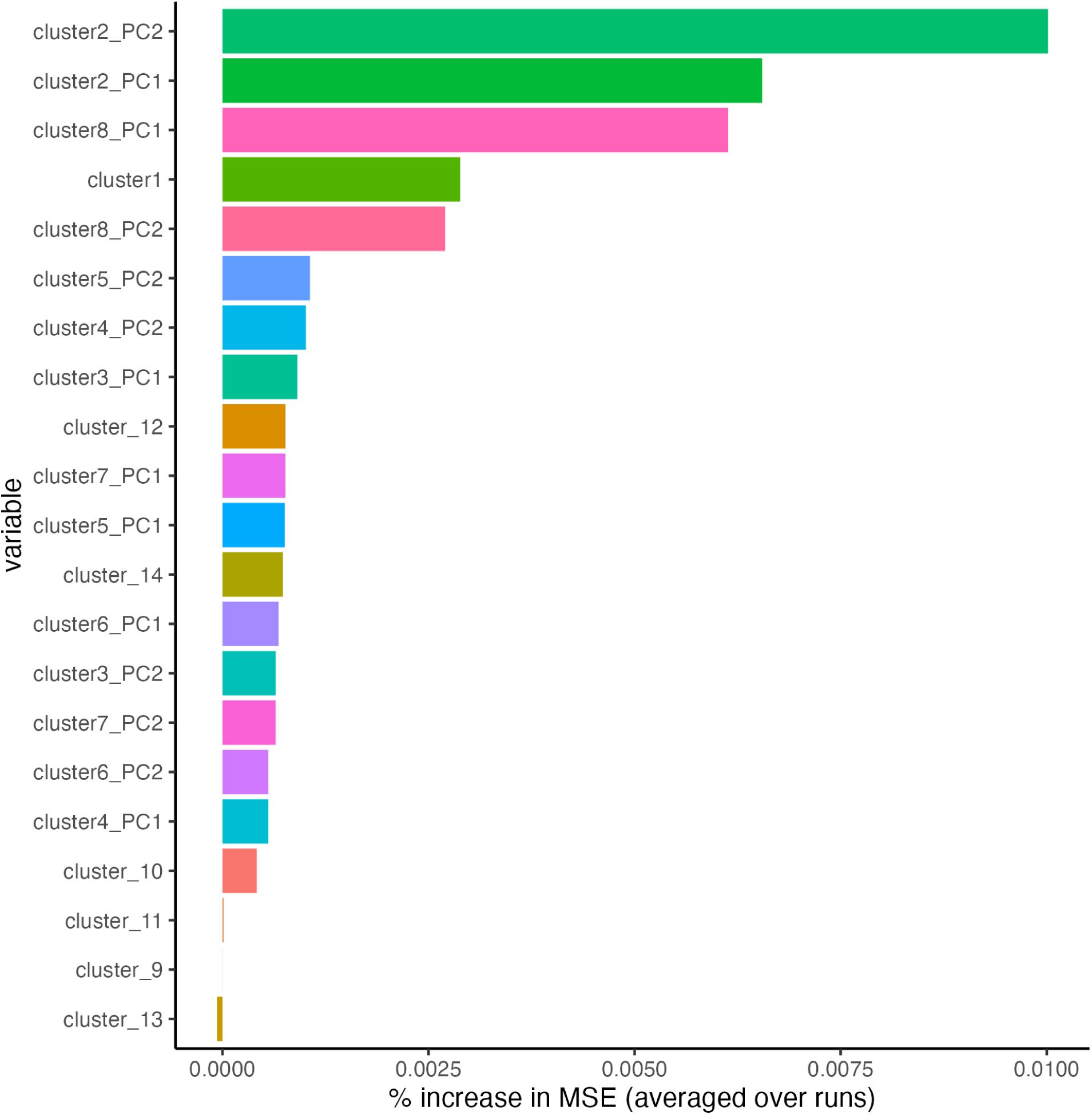
Estimated variable importance measures, i.e. % increase in mean squared error or MSE, from pre-grouped random forest implementation (RF #2) for the UK. Rows indicate cluster names (a full list of variables belonging to each cluster can be found in Supplementary Table S3) and corresponding principal components summarizing the information contained in the cluster, if the cluster consists of multiple variables. PC1 denotes principal component 1 and PC2 denotes principal component 2.

**Figure 2c.**
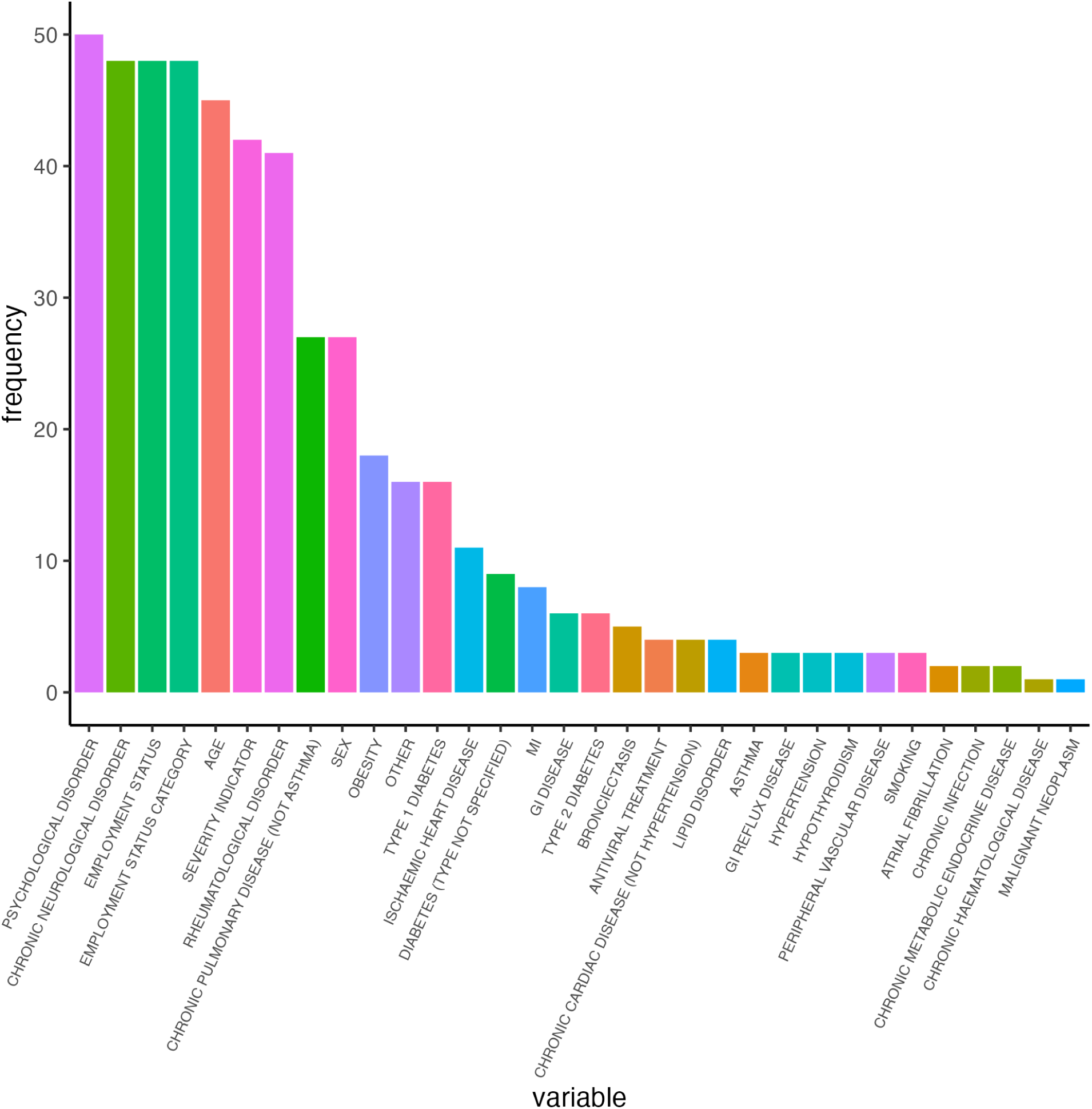
Number of times (frequency) each variable appears in clusters selected as most informative for each run of the model-grouped procedure (RF #3) for the UK.

Increased income/job stability, as proxied by employment status, was consistently associated with increased QoL utility scores, irrespective of the binary designation. We found that self-reported full-time employment versus any other employment status category (excluding retirement) was significantly associated with, on average, 0.0908 (0.0434, 0.138) increased units of QoL utility scores, via the direct effect, and 0.0139 (0.000804, 0.0271) increased units of QoL utility scores, via the indirect effect, with a proportion non-mediated of 0.894 (0.746, 0.996). We obtained an even stronger and significant relationship between self-reported full-time employment versus unemployment, with, on average, 0.216 (0.136, 0.297) increased QoL utility scores observed among the full-time employed relative to the unemployed (direct) and 0.0353 (0.0140, 0.0566) increased QoL utility scores observed among the full-time employed versus unemployed (indirect). The proportion non-mediated was 0.886 (0.804, 0.967), suggesting that 88.6% of the positive adjusted association between full-time employment versus unemployment and long COVID QoL utility scores does not operate through the considered intermediates.

Female sex was associated with lower expected QoL utility scores, with a direct and indirect effect of -0.0674 (-0.105, -0.0300) and -0.0247 (-0.0427, -0.00675), respectively. The corresponding proportion non-mediated was the lowest observed among all contrasts, with 67.6% (45.5%, 89.7%) of the negative adjusted association between female sex and long COVID QoL utility scores being unexplained by the clinical intermediates.

### Russia

The median (IQR) of long COVID QoL utility scores was 0.966 (0.916,1.0). Full-time employment (55%) and retirement (39%) were the predominant employment status categories. Long COVID QoL scores were highest among students, part-time employees, full-time employees, and carers, and lowest among those in the retired and unemployed categories. Males reported higher long COVID QOL utility scores than females (p<0.001). The mean age of participants was 59.6 years (SD: 14.4 years) and hypertension was the most frequently reported comorbidity (59%).

Age, followed by employment status indicators, hypertension, and chronic neurological disorder, outranked all other variables in predicting QoL utility scores in this cohort (Figure 3a). The pre-grouped approach generally supported these findings. The group containing solely age led the rankings; the groups containing the socio-demographic variables, hypertension and other cardiac disease, and dementia and chronic neurological disorder then followed (Figure 3b). Similarly, from the model-grouped approach, age, chronic neurological disorder, employment status indicators, hypertension and dementia led the set of most frequently selected variables (Figure 3c).

**Figure 3a.**
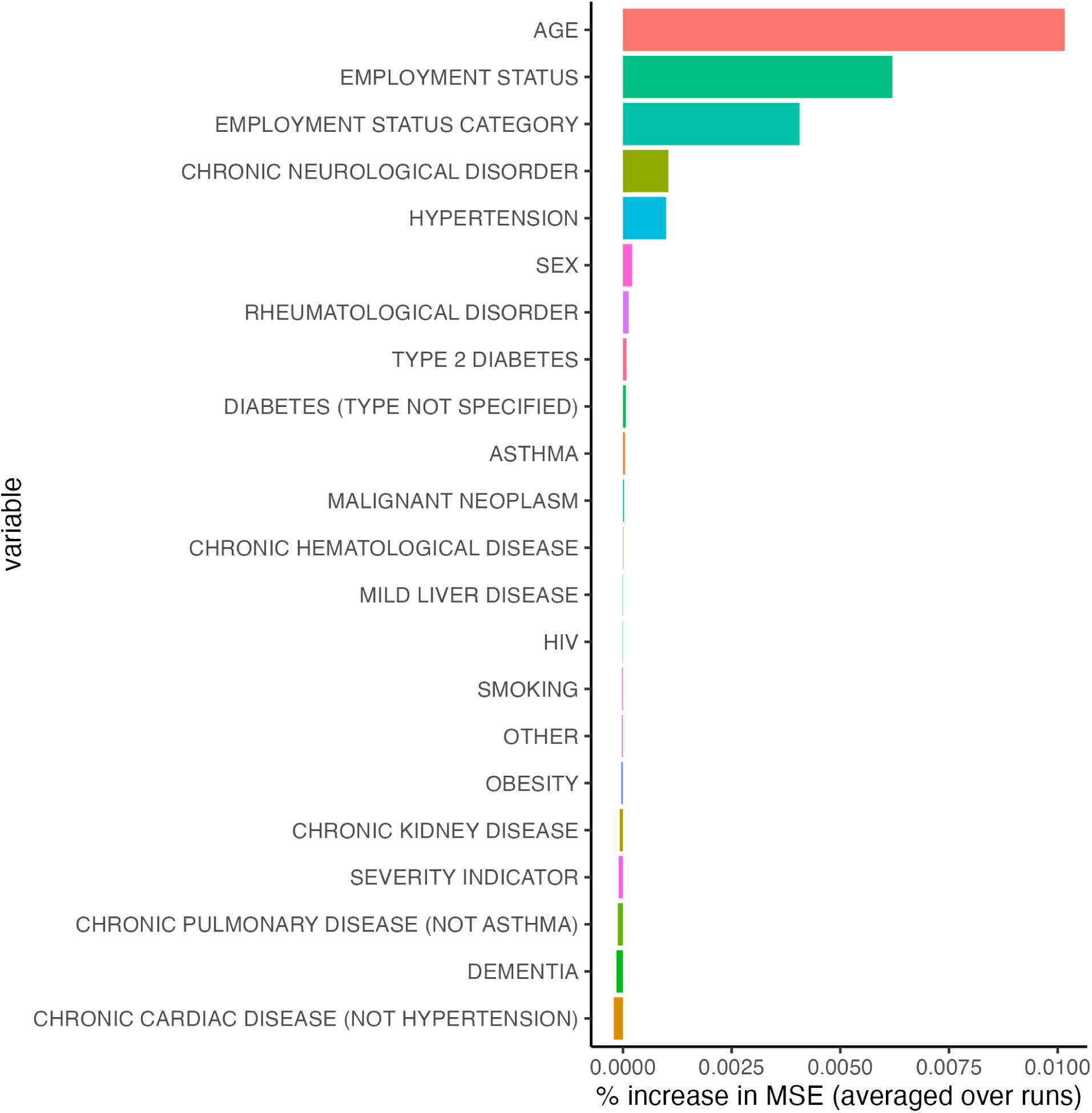
Estimated variable importance measures, i.e. % increase in mean squared error or MSE, from individual random forest implementation (RF #1) for Russia. Variables with negative % MSE values are considered unimportant.

**Figure 3b.**
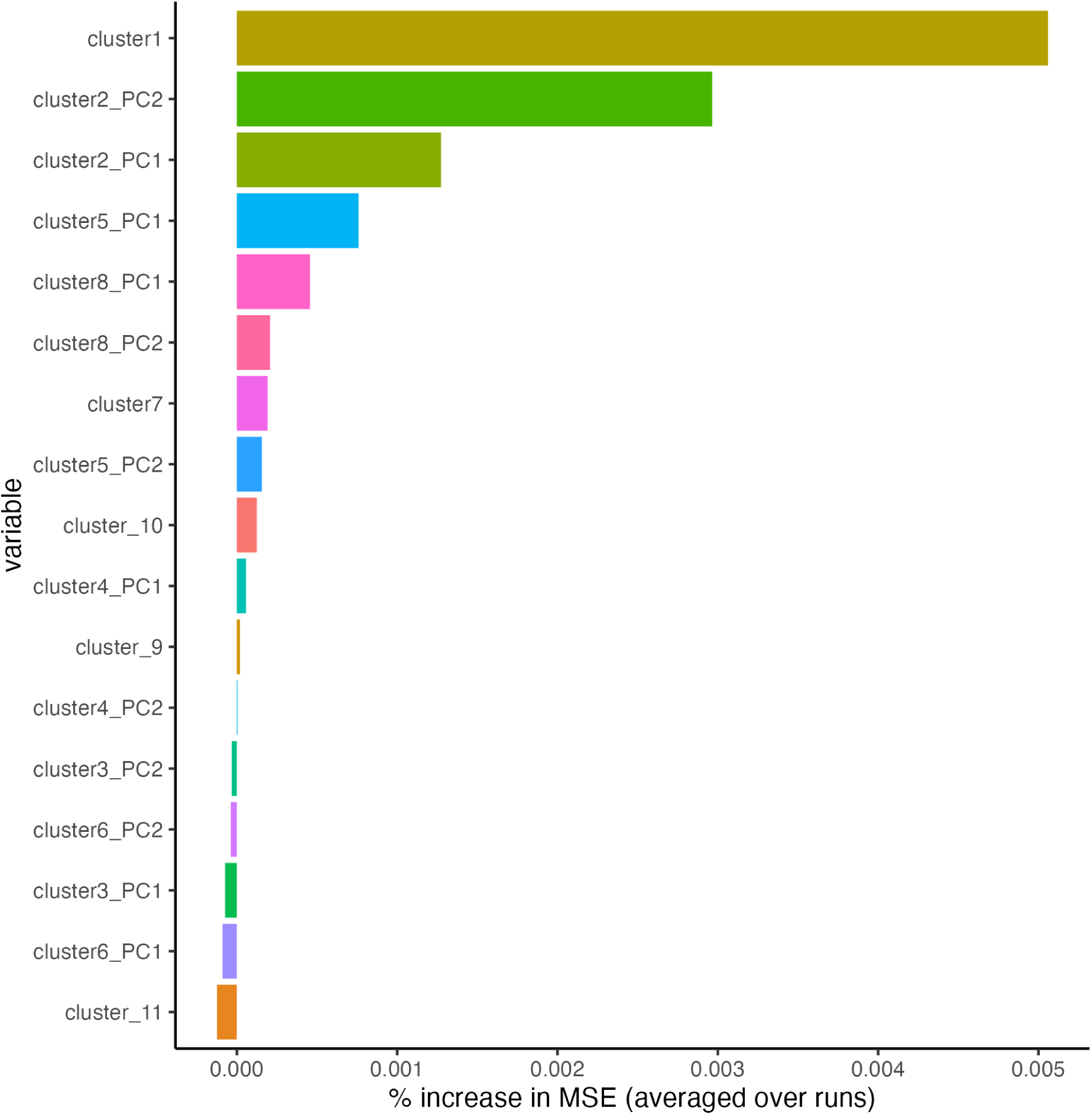
Estimated variable importance measures, i.e. % increase in mean squared error or MSE, from pre-grouped random forest implementation (RF #2) for Russia. Rows indicate cluster names (a full list of variables belonging to each cluster can be found in Supplementary Table S3) and corresponding principal components summarizing the information contained in the cluster, if the cluster consists of multiple variables. PC1 denotes principal component 1 and PC2 denotes principal component 2.

**Figure 3c.**
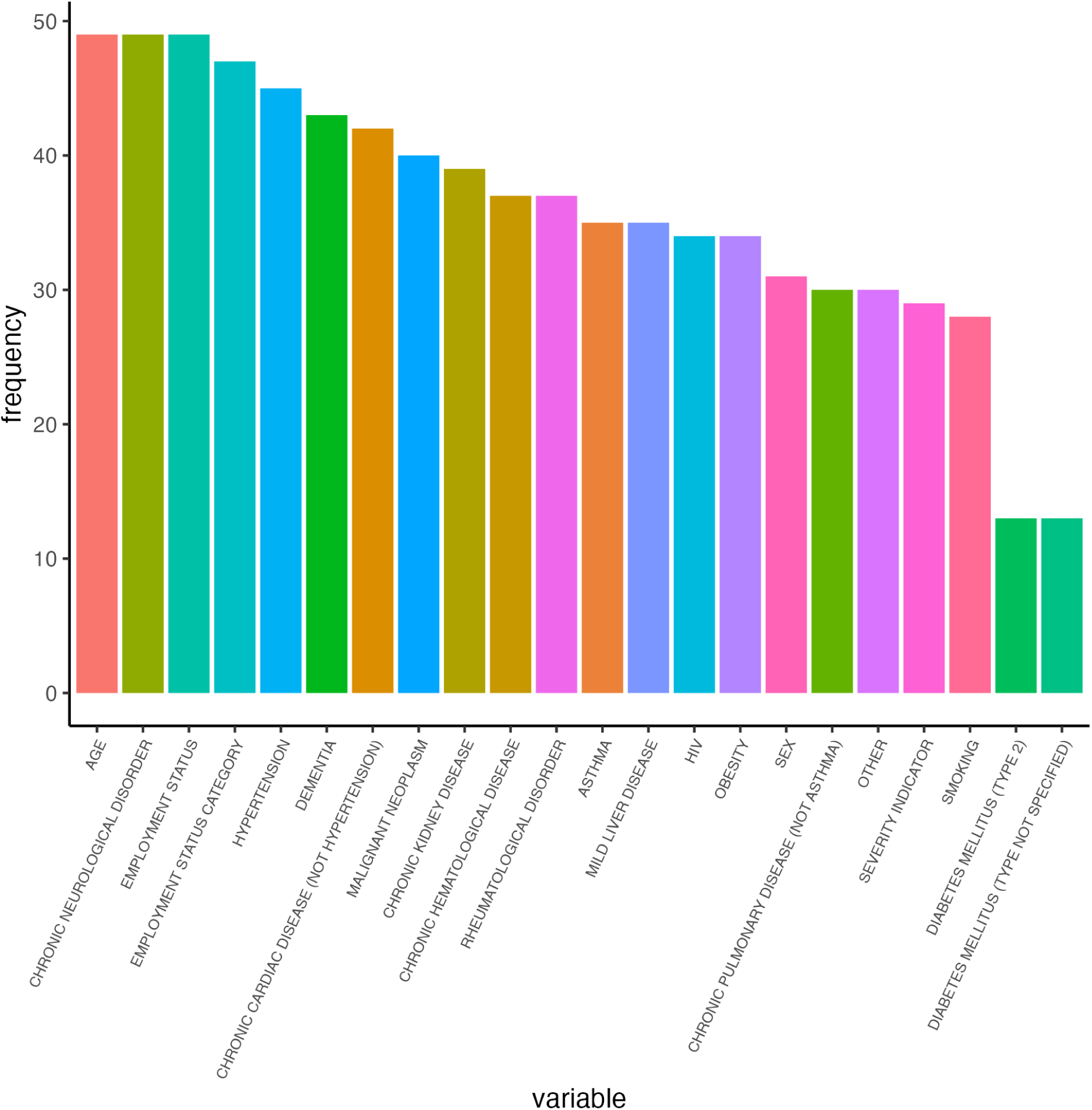
Number of times (frequency) each variable appears in clusters selected as most informative for each run of the model-grouped procedure (RF #3) for Russia.

Full-time employment was associated with higher long COVID QoL utility scores compared to all other employment status categories via the direct effect (0.02085, 95% CI: (0.00344, 0.0383)) but marginally lower long COVID QoL utility scores via the indirect effect (-0.000771, 95% CI: (-0.00384, 0.00229)), suggesting that clinical and non-clinical pathways operate in opposing directions in this cohort. As such, the estimated proportion mediated was negative, which corresponds to a proportion non-mediated exceeding 1. There were insufficient numbers in select other employment status categories in order to evaluate additional pairwise contrasts.

Female sex was associated with lower QoL utility scores, with an estimated direct effect of -0.0102 (-0.0187, -0.0017) and indirect effect of -0.00171 (-0.00420, 0.000783), corresponding to a proportion non-mediated of 0.872 (0.713, 1). That is, 87.2% of the adjusted association between female sex and long COVID QoL could not be ascribed to the considered clinical intermediates.

## Discussion

In this study, we provided a quantitative assessment of the extent to which social factors, compared to commonly highlighted clinical conditions, may relate to varying experiences with long COVID. We addressed this overarching question in two stages, first comparing the predictive role of clinical and social factors on long COVID QoL and subsequently distilling the unique contribution of social factors on differences in long COVID QoL, abstracting away intermediate comorbidity impacts.

In addition to neuropsychological and rheumatological comorbidities and age, we found that educational attainment or employment status, and to a lesser extent sex at birth, were generally as or more predictive of long COVID QoL utility scores. Our statistical mediation analyses further suggest that not only are indicators of social disadvantage notably predictive of lower QoL utility scores, i.e., poorer QoL, but also that the connection between these variables and QoL could only be partially explained by key long COVID-predicting comorbidities. This general finding, i.e., that disparities are not solely attributable to underlying differences in comorbidity rates across various demographic groups, has been validated in other studies conducted over the course of the COVID-19 pandemic (12,17,33).

Our study benefited from the use of a sizable and multi-national cohort with long-term QoL data, providing more nuanced information on post-acute COVID-19 experiences beyond simply whether patients experienced long COVID symptoms. Given the reasonably large sample sizes of our study cohorts, we were able to apply flexible data-adaptive machine learning tools, including recent developments in causal machine learning (30–32). The variable selection methods we used allow us to avoid relying on restrictive modelling assumptions and further accommodate inherent variable groupings. The statistical mediation approaches we applied integrate flexible algorithms to estimate nuisance parameters and semi-parametric theory for best-in-class estimation (30–32), providing an alternative to strictly parametric approaches (34) that place restrictive assumptions about the underlying and unknown data-generating process.

There are several important limitations of our analysis. First, as we noted, we did not have information on subject-specific duration of long COVID, so we could not evaluate a measure of long COVID-adjusted quality of life over the full time span of the condition. Additionally, we were unable to examine the varying roles of the different social and clinical factors on *changes* in QoL, due to the aforementioned issues with recall. However, to informally assess whether differences in post-COVID QoL between socioeconomic groups and self-reported sex were simply artefacts of baseline QoL differences, we compared pre-COVID QoL scores across groups. We found that for Norway and Russia, cross-group variability in estimated responses by EQ-5D-5L dimensions was generally higher post-versus pre-COVID, based on variability of medians by group by dimension, suggesting that pre-COVID heterogeneity in QoL did not fully drive the differences we observed across groups (Supplementary Appendix). The UK cohort had pre-COVID QoL measures for only 21% of subjects, so we could not make a standardised comparison. For future work, it is imperative to collect information on QoL measures at all stages of illness, not simply ex post facto, by positioning readily implementable study protocols at the outset of an outbreak, and tabulating information about duration of symptoms.

It can also be concluded that survivorship bias (35) might affect our results, as only subjects who completed a follow-up survey at any follow-up interval can have their QoL measures recorded, and those lost to follow up due to debilitating medical events are also likely to have a far reduced QoL prior to this event. However, we note that no participants in the three cohorts died at any point during follow-up. Furthermore, our study may be limited by the fact that it self-selects for subjects who were able to contact the healthcare system while they were ill with COVID-19, irrespective of disease severity, and thus misses those who were not seen in a hospital setting. For instance, we may not be effectively accounting for populations who have reduced overall healthcare access who would not have been able to visit a hospital setting during their illness. If such populations generally experienced lower QoL, then the estimates we provided here would provide a lower bound of the relationship between socioeconomic disadvantage and QoL with long COVID.

Additionally, we were limited to specific socio-economic variables that may not fully reflect participants’ levels of socio-economic deprivation, which further varied by country. Thus, we can only draw conclusions about the role of socio-economic status in relation to the specific measures defined for each country. For future work, it would be useful to emphasise the collection of more proximate *shared* indicators of socio-economic status. We were also limited to data on sex at birth, which does not capture important gender-based disparities that exist beyond this binary (36).

Finally, while there was a general consensus in the rankings of clinical and social factors and the direction and magnitudes of the association between social factors and long COVID utility scores, we did observe some variation in these results across the study settings. For instance, among clinical factors, neurological and rheumatological disorders were key identified predictors of long COVID QoL in the Norway and UK cohorts, while cardiological factors also played an important role in the Russia cohort. Additionally, we found differing directions for the direct and indirect effects of full-time employment status in the Russia cohort, which we did not observe for related contrasts in the other two cohorts. Such heterogeneity is expected across study settings, each of which consist of unique social and structural systems, cultures, and patterns of healthcare access, and points to the importance of drawing setting-specific conclusions about the role of a given social factor on long COVID QoL.

### Conclusion

Here, we apply a robust statistical framework to highlight the role of social disparities in chronic ill-health. Our data highlights the multifactorial relationship between pre-existing risk factors and socio-economic factors and long COVID QoL. As such, we demonstrate that accounting for social vulnerabilities when evaluating determinants of post-acute COVID-19 trajectories is essential and that programmes focusing solely on clinical targets may not be sufficient. Conversely, transformational societal interventions, which address access to care, education, employment, etc., have the opportunity to lead to potentially more comprehensive benefits and improve overall well-being in marginalised communities.

## ISARIC Clinical Characterisation Group

Beatrice Alex, Eyvind W. Axelsen, Benjamin Bach, John Kenneth Baillie, Wendy S. Barclay, Joaquín Baruch, Husna Begum, Lucille Blumberg, Debby Bogaert, Fernando Augusto Bozza, Sonja Hjellegjerde Brunvoll, Polina Bugaeva, Aidan Burrell, Denis Butnaru, Roar Bævre-Jensen, Gail Carson, Meera Chand, Barbara Wanjiru Citarella, Sara Clohisey, Marie Connor, Graham S. Cooke, Andrew Dagens, John Arne Dahl, Jo Dalton, Ana da Silva Filipe, Emmanuelle Denis, Thushan de Silva, Pathik Dhangar, Annemarie B. Docherty, Christl A. Donnelly, Thomas Drake, Murray Dryden, Susanne Dudman, Jake Dunning, Anne Margarita Dyrhol-Riise, Linn Margrete Eggesbø, Merete Ellingjord-Dale, Cameron J. Fairfield, Tom Fletcher, Victor Fomin, Robert A. Fowler, Christophe Fraser, Linda Gail Skeie, Carrol Gamble, Michelle Girvan, Petr Glybochko, Christopher A. Green, William Greenhalf, Fiona Griffiths, Matthew Hall, Sophie Halpin, Bato Hammarström, Hayley Hardwick, Ewen M. Harrison, Janet Harrison, Lars Heggelund, Ross Hendry, Rupert Higgins, Antonia Ho, Jan Cato Holter, Peter Horby, Samreen Ijaz, Mette Stausland Istre, Clare Jackson, Waasila Jassat, Synne Jenum, Silje Bakken Jørgensen, Karl Trygve Kalleberg, Christiana Kartsonaki, Seán Keating, Sadie Kelly, Kalynn Kennon, Saye Khoo, Beathe Kiland Granerud, Anders Benjamin Kildal, Eyrun Floerecke Kjetland, Paul Klenerman, Gry Kloumann Bekken, Stephen R Knight, Andy Law, Jennifer Lee, Gary Leeming, Wei Shen Lim, Andreas Lind, Miles Lunn, Laura Marsh, John Marshall, Colin McArthur, Sarah E. McDonald, Kenneth A. McLean, Alexander J. Mentzer, Laura Merson, Alison M. Meynert, Sarah Moore, Shona C. Moore, Caroline Mudara, Daniel Munblit, Srinivas Murthy, Fredrik Müller, Karl Erik Müller, Nikita Nekliudov, Alistair D Nichol, Mahdad Noursadeghi, Anders Benteson Nygaard, Piero L. Olliaro, Wilna Oosthuyzen, Peter Openshaw, Massimo Palmarini, Carlo Palmieri, Prasan Kumar Panda, Rachael Parke, William A. Paxton, Frank Olav Pettersen, Riinu Pius, Georgios Pollakis, Mark G. Pritchard, Else Quist-Paulsen, Dag Henrik Reikvam, David L. Robertson, Amanda Rojek, Clark D. Russell, Aleksander Rygh Holten, Vanessa Sancho-Shimizu, Egle Saviciute, Janet T. Scott, Malcolm G. Semple, Catherine A. Shaw, Victoria Shaw, Louise Sigfrid, Mahendra Singh, Vegard Skogen, Sue Smith, Lene Bergendal Solberg, Tom Solomon, Shiranee Sriskandan, Trude Steinsvik, Birgitte Stiksrud, David Stuart, Charlotte Summers, Andrey Svistunov, Arne Søraas, Emma C. Thomson, Mathew Thorpe, Ryan S. Thwaites, Peter S Timashev, Kristian Tonby, Lance C.W. Turtle, Anders Tveita, Timothy M. Uyeki, Steve Webb, Jia Wei, Murray Wham, Maria Zambon.

## Acknowledgments

The investigators thank all the clinical and research staff, who performed the follow-up assessments and collected this data, and the participants for their individual contributions in these difficult times. We would also like to thank the Long Covid Support group and ISARIC’s Global Support Centre for their invaluable support.

We also acknowledge the support of the COVID clinical management team, AIIMS, Rishikesh, India; the Liverpool School of Tropical Medicine and the University of Oxford; Imperial NIHR Biomedical Research Centre; the dedication and hard work of the Norwegian SARS-CoV-2 study team; and preparedness work conducted by the Short Period Incidence Study of Severe Acute Respiratory Infection.

This work uses data provided by patients and collected by the NHS as part of their care and support #DataSavesLives. The data used for this research were obtained from ISARIC4C. We are extremely grateful to the 2648 frontline NHS clinical and research staff and volunteer medical students who collected these data in challenging circumstances; and the generosity of the patients and their families for their individual contributions in these difficult times. The COVID-19 Clinical Information Network (CO-CIN) data was collated by ISARIC4C Investigators. We also acknowledge the support of Jeremy J Farrar and Nahoko Shindo.

*The computations in this paper were run on the FASRC Cannon cluster supported by the FAS Division of Science Research Computing Group at Harvard University*

