## Supplementary Appendix for "Modelling the relative influence of socio-demographic variables on post-acute COVID-19 quality of life"

### Supplementary methods

A. Long COVID utility score calculation

Long COVID utility scores were computed as the complement of the sum of the relevant utility weights for each participant

$$Long\ COVID\ utility_i = (1 - \sum_{k=1}^{5} QALY.weight.post_{ki})$$

Where i denotes subject, k denotes each of the five items of the EQ-5D-5L survey, $QALY.weight.post_{ki}$ is the weight from the appropriate value set corresponding to the chosen response for item k for subject i.

B. Variable preparation, missing data imputation, and post-stratification weighting

In the UK and Russian cohorts, COVID-19 disease severity was defined following the categorization in Reyes et al. (1). That is, subjects who reported any of the following outcomes (high-flow nasal cannula, ventilation, or the use of inotropes or vasopressors) during their hospital admission were classified as having severe acute COVID-19 disease (1). Severity indicators were classified as unknown among subjects who had missing entries for all five possible indicators. In the Norway cohort, there was little information available on any of the aforementioned outcomes and another measure of acute COVID-19 severity, hospitalization for acute COVID-19 infection (any or ICU admission), was very sparsely recorded. Therefore, we excluded this variable among our factors under consideration in this cohort.

Vaccination data was obtained using a search for the following key terms within the field for immunization against COVID-19: Moderna, Pfizer, AstraZeneca, and Janssen, as well as “COVID vaccination”, “COVID-19 vaccination”, and “COVID-19 vaccine type”, which were the general categories recorded when no specific brand was noted.

Antiviral and additional treatment data was obtained using a search for the following key terms within the field for treatment: darunavir, remdesivir, acyclovir, valganciclovir, lopinavir/ritonavir, metformin, as well as “Antiviral agent”, which was a general category recorded when no specific type was noted. We selected these drugs as those explicitly listed as possible options in the ISARIC follow-up surveys, as well as a diabetes drug that has been identified to be protective against long COVID (metformin) (2). Data on the two most currently supported therapies nirmatrelvir and molnupiravir was not available in any cohort.

Missingness was reported for the following variables: Norway: educational attainment (17.6%), race/ethnicity (2.3%), sex (1.2%), age (0.006%), vaccination status (0.90%), antiviral treatment during illness (83.5%), and the severity indicator (75.7%); UK: employment status (11.2%), race/ethnicity (1.6%), vaccination status (66.5%), and antiviral treatment during illness (22.3%); and Russia: employment status (4.8%), race/ethnicity (100%), vaccination status (100%), and antiviral treatment during illness (100%).

Missing variable imputation was performed via Multiple Imputation by Chained Equations (MICE) using the *mice* package in R (3). We cannot make a definitive conclusion on the missingness mechanisms for sex at birth . That is, we have no reason to believe that females compared to males are less likely to self-report their sex, i.e. we do not have any evidence in favor of a significant difference or missing not at random (MNAR) mechanism. Thus, the use of MICE, which assumes a missing (completely) at random mechanism, appears justified. However, vaccination status and the socio-economic indicators of educational attainment and employment status are more likely to be MNAR due to social stigmas and fear of judgment. Nonetheless, a relatively low fraction of most of these values were missing, with the exception of the vaccination status indicator in the UK and Russia cohorts, antiviral treatment in the Norway and Russia cohorts, and the severity indicator in the Norway cohort. Consequently, to avoid imposing significant bias, we chose not to impute these specific variables and excluded them from their respective variable sets. For all cohorts, we specified 10 imputations and 10 iterations following standard practice, where, for each iteration, missing variables were expressed as a function of all other variables via polytomous regression (for categorical variables), logistic regression (for binary variables), and predictive mean matching (for continuous variables). Missing values were then assigned the majority imputation across runs for the corresponding variable.

The distributions of sex at birth in our sampled populations non-negligibly deviated from those in the underlying Norwegian and British populations (4,5). Thus, to correct for this binary mis-representativeness, we conducted sensitivity analyses where subjects were assigned post-stratification weights according to the *raking* method (6–8), applying the *anesrake* package in R (9). In brief, this approach adjusts the distribution of each of the sampled populations on the basis of sex to correspond to its distribution in the overall population (6–8).

C. Quantifying variable importance

For all analyses, comorbidities were filtered to only include conditions with at least ten subjects reporting having them. All of our random forest regressions were trained on 80% of the dataset using the *caret* package (10). For the individual RF and pre-grouped RF, variables were ranked according to their associated % increase in mean squared error, averaged over 100 runs. To run these RFs, we used the *randomForest* package (11). While we averaged our results across 100 runs for the individual and pre-grouped analyses, we considered a fewer number of runs (50) for the model-grouped analysis to avoid exorbitant model costs, due to the resource intensiveness of each CoV-VSURF run, using the *CoVVSURF* package (12). For Norway, we used dummy variables representing belonging to each quintile of educational attainment compared to the referent category of quintile 1 for RF #3, as CoV-VSURF does not accept ordered factors as inputs. Finally, for the population adjustment sensitivity analysis, weights were incorporated in all random forest implementations using the sample_weight parameter in the randomForest (11) and covsurf (12) functions.

For our pre-grouped random forest regression implementation, we applied multiple factor analysis to each cluster, which corresponds to a principal component analysis for numeric variables and multiple correspondence analysis for categorical variables, considering the first and second principal components of each. For binary variables, we replaced subjects' values with the loading obtained for that variable, with variable coordinates defined as the square root of the component's eigenvalue (13), if present, and with 0, if absent. For categorical variables with more than one level, we replaced subjects' values with the loading obtained for the relevant level of that variable. For numeric variables, we multiplied subjects' values by the loading obtained for that variable. For each cluster, we then summed the resulting weighted variables by subject and by component, reducing the number of variables for each cluster.i from $p_{cluster.i}$ to two. The resulting reduced number of covariates were then used as features for our random forest model runs. We used the *FactoMineR* package (14) to conduct this cluster summary analysis.

D. Estimating NDEs, NIEs, and proportions non-mediated

*Defining exposures*

For Norway, our binary proxy of SES was high/low educational attainment, where 'high' encompassed quintiles 3-5 and 'low' encompassed quintiles 1 and 2. For the UK and Russia, our binary proxy of SES was high/low employment status. For the UK, 'high' encompassed full-time employment and 'low' encompassed all other employment status categories (furloughed, part-time employment, student, and the unemployed), with the exception of retirement. For Russia, 'high' encompassed full-time employment and 'low' encompassed all other employment status categories, where retirees are included due to notable disparities in representation of full-time employees versus all other categories in this cohort.

*Defining mediators*

Mediators were selected following a literature search of studies distinguishing key long COVID predictors (15,16), consistent with the data available for each country, resulting in the following final mediator sets: asthma, chronic cardiac disease (not hypertension), hypertension, chronic pulmonary disease (not asthma), type 1 diabetes (T1D), type 2 diabetes (T2D), diabetes (type not specified), psychological disorder, smoking, and vaccination status (Norway), asthma, chronic cardiac disease (not hypertension), hypertension, chronic pulmonary disease (not asthma), T2D, diabetes (type not specified), psychological disorder, ischemic heart disease, smoking, obesity, and antiviral treatment (UK) and asthma, chronic cardiac disease (not hypertension), hypertension, chronic pulmonary disease (not asthma), T2D, diabetes (type not specified), smoking, and obesity (Russia).

*Running mediation analyses*

Estimating the NDE, NIE, and proportion mediated via targeted maximum likelihood estimation (TMLE) requires two key steps (17,18). First, the data is divided into $k$ cross-validation folds (subsets of the data), to which we train and test our multiple algorithms of choice, where the response measure is the expected mean of long COVID QoL utility scores dependent on each of our binary SES proxies or female sex, the confounders, and mediators (17,18). The expected means of long COVID QoL utility scores dependent on each of our binary SES proxies or female sex, the confounders, and mediators under the two levels of the exposure are then estimated using the weighted sum of each algorithm's fit to the original data (17,18). The difference in these fitted means of long COVID QoL utility scores dependent on each of our binary SES proxies or female sex, the confounders, and mediators for each level of the exposure, alongside the conditional probability of each of our binary SES proxies or female sex, dependent on confounders and the conditional probability of each of our binary SES proxies or female sex, dependent on confounders and mediators are subsequently used as ingredients to compute estimates of the NDE and NIE (17,18). For the population adjustment sensitivity analysis, weights were incorporated using the *survey_weight* parameter in the *medoutcon* package (19).

To estimate the required parameters we incorporated the following parametric methods: a simple intercept regression model and generalized additive models (GAMs) (20), the latter equipped to accommodate non-linear forms for continuous covariates, as well as nonparametric gradient boosted regression trees (21), applying the *lightgbm* package (22), and feed-forward neural networks (23), applying the *nnet* package (24), both of which can better capture underlying nonlinear trends non-parametrically. As noted, algorithms are combined via a super learner, applying the *sl3* package (25), through a non-negative least squares meta-learner, applying the *nnls* package (26). All COVID utility scores were normalized to fall within a range of [0,1] prior to being used as inputs in the super learner. Note that the exact relationship between the NDE/NIE and proportion non-mediated may not hold for numerical reasons, as point estimates and CIs are estimated using cross-validation and separately for each of the measures.

Directed acyclic graphs (DAGs) were drawn using the *dagitty* (27) and *ggdag* (28) packages.

*Required assumptions for interpreting associational measures causally*

Formally, let Z capture the baseline confounders (sex and age for the high SES contrast, and SES indicator for the female sex contrast), A captures the binary exposure category (high SES or female sex), M captures the considered mediators, and Y is the outcome (long COVD utility scores). Below we list the identifiability assumptions that would be required to supply our measures a causal interpretation:

- Exposure and mediator positivity: $P(A=a|Z=z)>0$ and $P(M=m|A=a,Z=z)>0$, i.e., there must exist at least one individual within each of the two SES categories and each of the two female sex categories, given baseline confounders and there must exist at least one individual with or without each mediator, given baseline confounders and SES or female sex level.
- Well-defined exposures and potential outcomes (consistency): If A=a, Y=**Y(a)**, i.e., the observed long COVID utility score for an individual with SES or female sex level = a must be equivalent to their counterfactual long COVID utility score under SES or female sex level = a. If high/low SES or female/male sex is not well-defined, then the observed outcomes under these values may vary on a per-person basis and thus may not be aligned with a consistent, well-defined counterfactual outcome.
- No interference between study units: An individual's SES or sex must not affect another individual's long COVID utility scores.
- No unmeasured confounding:
    - $\mathbf{Y(a,m) \perp A \mid Z}$ – no unmeasured confounding of the relationship between SES or female sex and long COVID utility scores, i.e., the counterfactual

outcome under SES or female sex = a and mediators = m is independent of the SES or female sex observed, conditional on baseline confounders.

- **M(a) $\perp$ A | Z** – no unmeasured confounding of the relationship between the mediators and SES or female sex, i.e., the counterfactual mediator under SES or female sex = a is independent of the SES or female sex observed, conditional on baseline confounders.
- **Y(a,m) $\perp$ M | Z,A** – no unmeasured confounding of the relationship between long COVID utility scores and the mediators, i.e., the counterfactual outcome under SES or female sex = a and mediators = m is independent of the mediators observed, conditional on baseline confounders and SES or sex.

E. Additional analyses

To compute the crude association between long COVID QoL utility scores and sex (female vs male), we conducted a one-sided Wilcoxon rank sum test which assumes independence and equal variance for the two groups, but does not assume normality of long COVID QoL utility scores.

All analyses were run using the R programming language and environment for statistical computing and graphics (version 4.2.2) (29).

### Supplementary results

*Comparison of EQ-5D-5L responses by demographic groups (in the final study populations for each cohort, post-missing data imputation)*

A. Norway

We found minimal differences across educational attainment quintiles in reported post-COVID responses for the dimensions anxiety/depression, mobility, self-care, and usual activities (Supplementary Figure S3A ii). However, quintiles 3-5 skewed towards lower responses for pain/discomfort, indicating fewer challenges for that dimension (Supplementary Figure S3A ii). Males and females reported similar distributions in responses for the EQ-5D-5L dimensions anxiety/depression, mobility, self-care, and usual activities (Supplementary Figure S4A ii). However, males skewed towards lower values for pain/discomfort (Supplementary Figure S4A ii).

Pre-COVID responses within each dimension were less variable across educational attainment groups compared to post-COVID responses (variance of medians = 0 for all categories vs 0 for all categories usual activities (0.2), pre- and post-COVID, respectively) (Supplementary Figure S3A i). Pre-COVID & post-COVID response maintained similar minimal variability across sex groups by dimension (variance of medians = 0 for all categories, pre- and post-COVID) (Supplementary Figure S4A i).

B. UK

There was considerable heterogeneity across employment status categories in responses to each of the five EQ-5D-5L dimensions (Supplementary Figure S3B).

Males and females reported similar distributions in responses for the EQ-5D-5L dimensions for mobility, pain/discomfort, and usual activities (Supplementary Figure S4B). However, males skewed towards lower values for anxiety/depression and self-care (Supplementary Figure S4B).

C. Russia

As with the UK, there was considerable heterogeneity across employment status categories in responses to each of the five EQ-5D-5L dimensions (Supplementary Figure S3C ii). Variability in responses was much lower for pre-COVID responses (Supplementary Figure S3C i) (variance of medians = 0 for all categories vs 0 for all categories except mobility (0.267) and pain/discomfort (0.175), pre- and post-COVID).

The distribution of EQ-5D-5L responses was nearly equivalent for both sexes on every dimension except mobility, where females skewed towards greater challenges (Supplementary Figure S4C ii). The same pattern was observed, with reduced variability for pre-COVID responses (Supplementary Figure S4C i) (variance of medians = 0 for all categories vs 0 for all categories and 0.5 for mobility for mobility, pre- and post-COVID).

*Sex-based population correction sensitivity analysis*

For both the Norway and UK cohorts, we observed consistent directionality and magnitude of estimates of the NDE, NIE, and proportion non-mediated, but with greater variability, in the population correction-sensitivity analysis for both the high SES and female sex contrasts (Supplementary Figures S5B and D and S6B and D). We also obtained a similar ranking of individual variables and grouped variables from all random forest implementations for the population correction-based sensitivity analysis (Supplementary Figures S8 and S9).

*Tables*

**Supplementary Table S1. Selected studies for extracting utility weights**

| Cohort | Study (Country) |
|---|---|
| Norway | Sun et al., 2022 (30) (Sweden) |
| UK | Devlin et al., 2018 (31) (England) |
| Russian cohort | Golicki et al., 2019 (32) (Poland) |

**Supplementary Table S2. Final employment status groupings**

| Group category | Norway | Russia |
|---|---|---|
| Retired | “Medically retired”, “Retired”, “Retired_Medically retired”, “Unable to work due to chronic illness_Retired” | “Early Retirement Due to Illness”, “Retired” |
| Unemployed | “Unemployed”, “Unable to work due to chronic illness” | “Unemployed”, “Unable to Work Due to Chronic Illness” |
| Full-time employment | “Full-time employment”, “Full time carer (children or other)”, “Full-time employment_Prefer not to say”, “Full-time employment_Full time carer (children or other)”, “Working full-time”, “Working Full-time' | “Working Full-Time” |
| Carer | NA (small n, so grouped w/ full-time employment above) | “Full Time Carer (Children or Others)”, |
| Furloughed | “Furloughed”, “Full-time employment_Furloughed” | NA |
| Student | “Student” | Student |
| Part-time employment | “Part-time employment”, “Working Part-time”, “Working part-time” | “Working Part-Time” |

**Supplementary Table S3. Predetermined groupings of variables, by country, for RF #2**

| Group | Norway | UK | Russia |
|---|---|---|---|
| 1 | Age | Age | Age |
| 2 | Educational attainment (years), educational attainment quintile, sex | Employment status, sex | Employment status, sex |
| 3 | Asthma, chronic pulmonary | Asthma, chronic pulmonary | Asthma, chronic |

| | | | |
|---|---|---|---|
| | disease (not asthma), smoking | disease (not asthma), bronchiectasis, smoking | pulmonary disease (not asthma), smoking |
| 4 | Obesity, type 1 diabetes, type 2 diabetes, diabetes (type not specified) | Obesity, type 1 diabetes, type 2 diabetes, diabetes (type not specified) | Obesity, type 2 diabetes, diabetes (type not specified) |
| 5 | Chronic cardiac disease (not hypertension), hypertension | Congestive heart failure, ischemic heart disease, atrial fibrillation, chronic cardiac disease (not hypertension), hypertension, peripheral vascular disease | Chronic cardiac disease (not hypertension), hypertension |
| 6 | Chronic kidney disease, liver disease (severity not specified) | Gastrointestinal (GI) disease, chronic metabolic endocrine disease, GI reflux disease, hypothyroidism, lipid disorder | Chronic kidney disease, mild liver disease |
| 7 | Chronic hematological disease, rheumatological disorder | Chronic hematological disease, rheumatological disorder | Rheumatological disorder |
| 8 | Psychological disorder, chronic neurological disorder | Psychological disorder, chronic neurological disorder | Dementia, chronic neurological disorder |
| 9 | Malignant neoplasm | Malignant neoplasm | Malignant neoplasm |
| 10 | Other | Myocardial infarction (MI) | Other |
| 11 | Vaccination status | Chronic infection | Country |
| 12 | N/A | Other | COVID-19 disease severity indicator |
| 13 | N/A | Antiviral treatment | N/A |
| 14 | N/A | COVID-19 disease severity indicator | N/A |

(A)

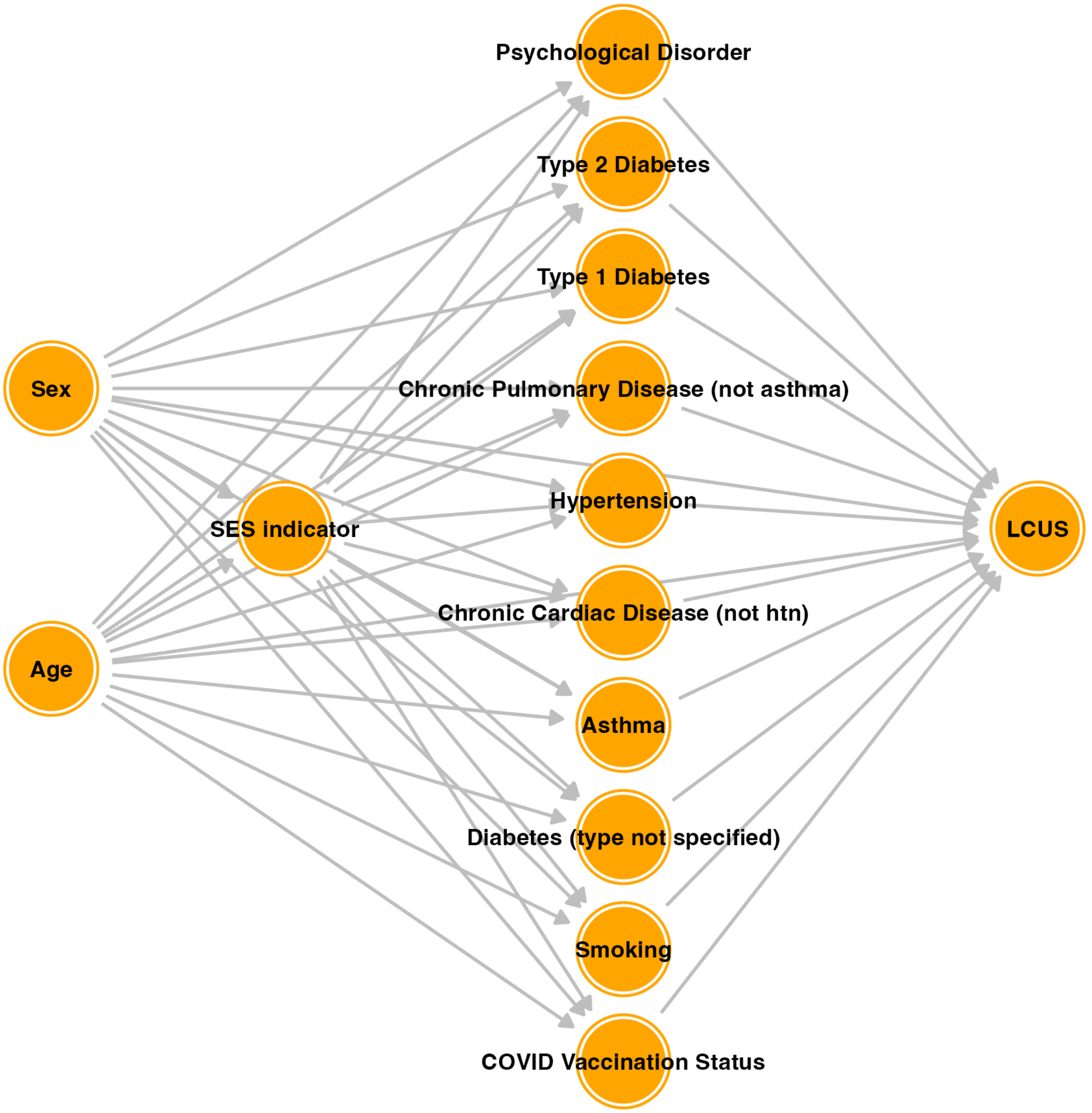
Psychological Disorder
Type 2 Diabetes
Type 1 Diabetes
Sex
Chronic Pulmonary Disease (not asthma)
SES indicator
Hypertension
LCUS
Chronic Cardiac Disease (not htn)
Age
Asthma
Diabetes (type not specified)
Smoking
COVID Vaccination Status

**(B)**

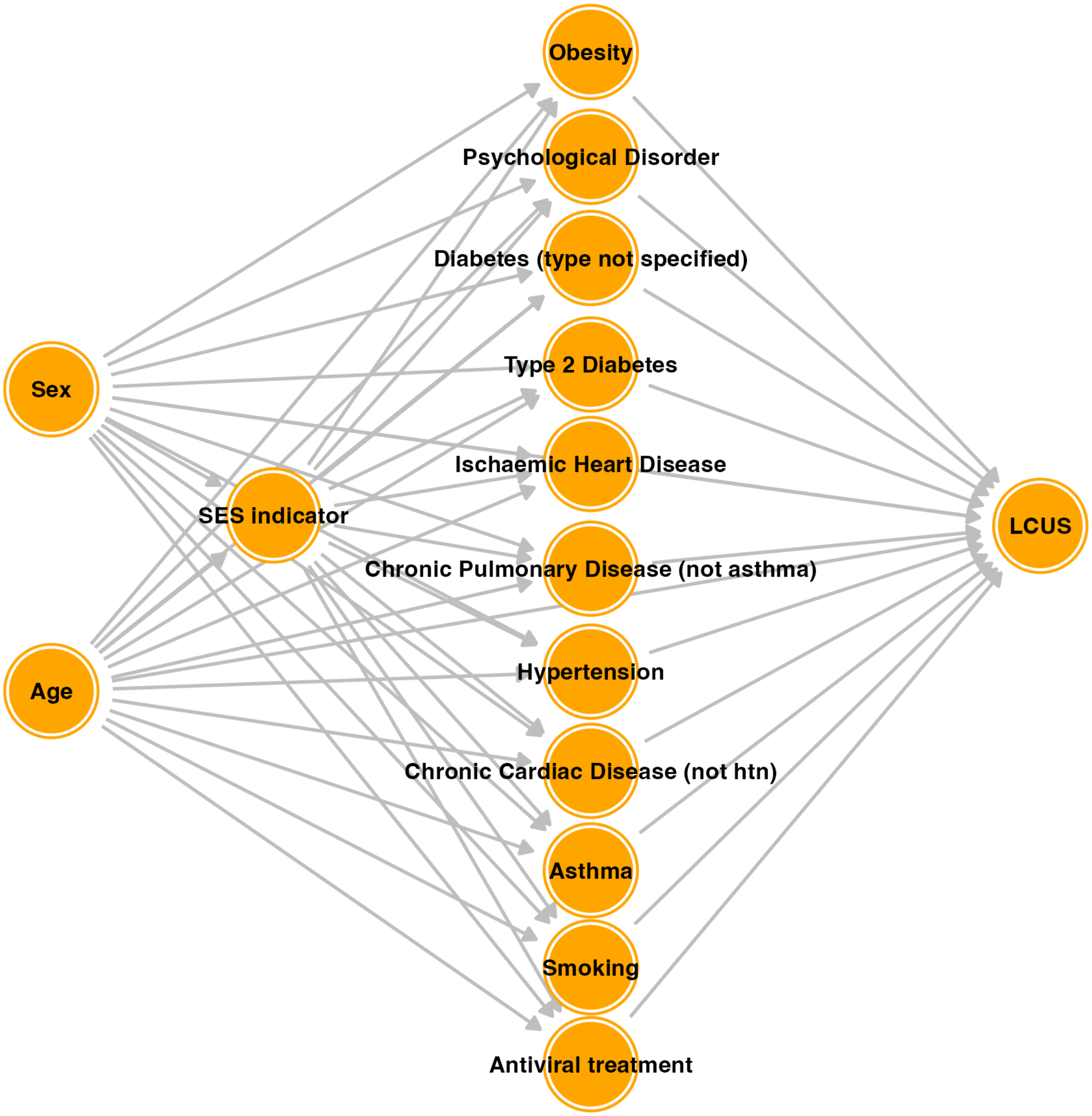
Obesity
Psychological Disorder
Diabetes (type not specified)
Type 2 Diabetes
Sex
Ischaemic Heart Disease
SES indicator
LCUS
Chronic Pulmonary Disease (not asthma)
Age
Hypertension
Chronic Cardiac Disease (not htn)
Asthma
Smoking
Antiviral treatment

**(C)**

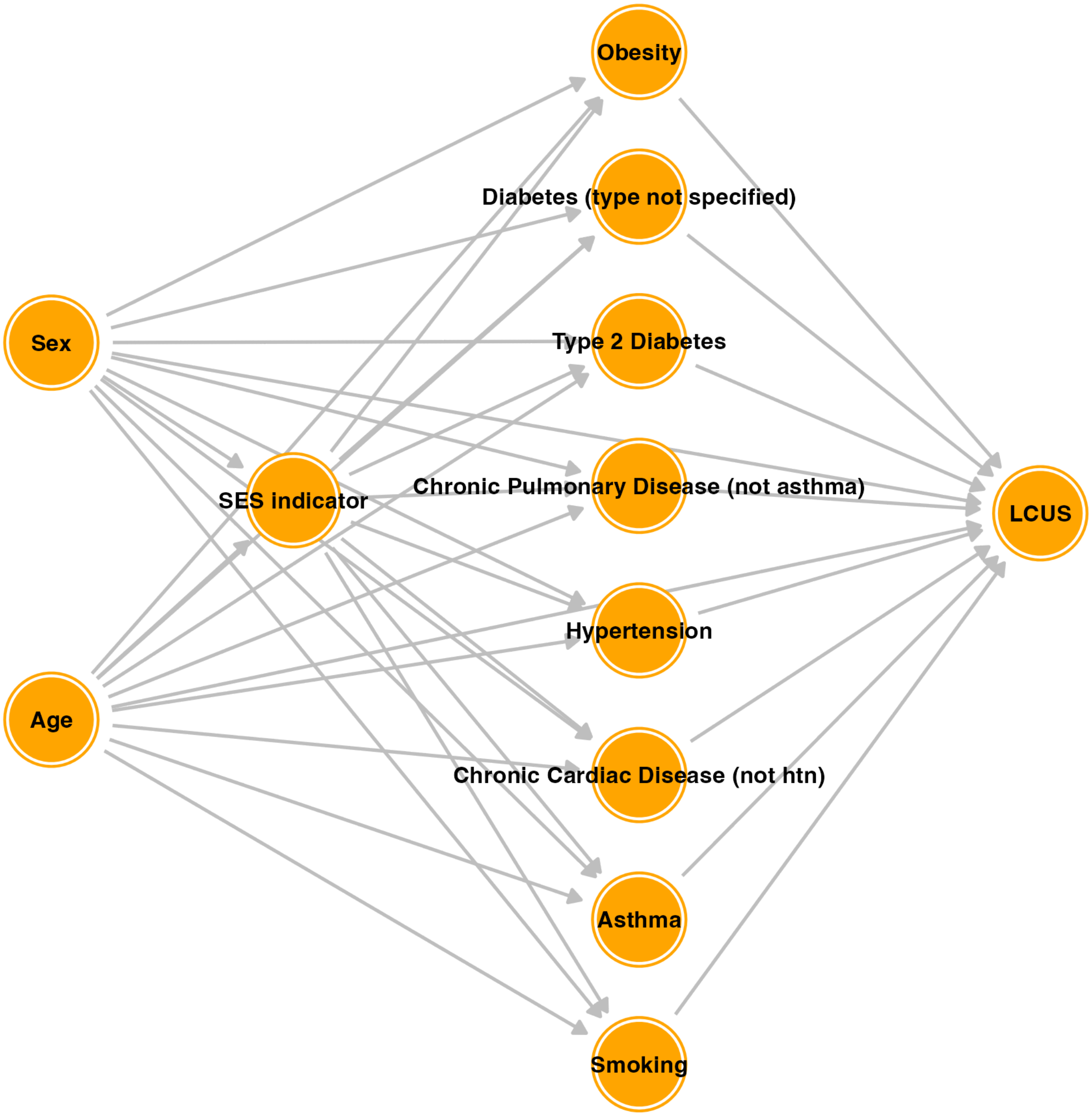


**Supplementary Figure S1. Assumed DAGs for the Norway, UK, and Russia cohorts with SES indicators as exposure. DAG for educational attainment binary variable in Norway (A), DAG for employment status binary variable in the UK (B), and DAG for employment status binary variable in the Russia cohort (C). Note: htn = hypertension, in "Chronic Cardiac Disease (not htn)." LCUS denotes Long COVID utility scores.**

(A)

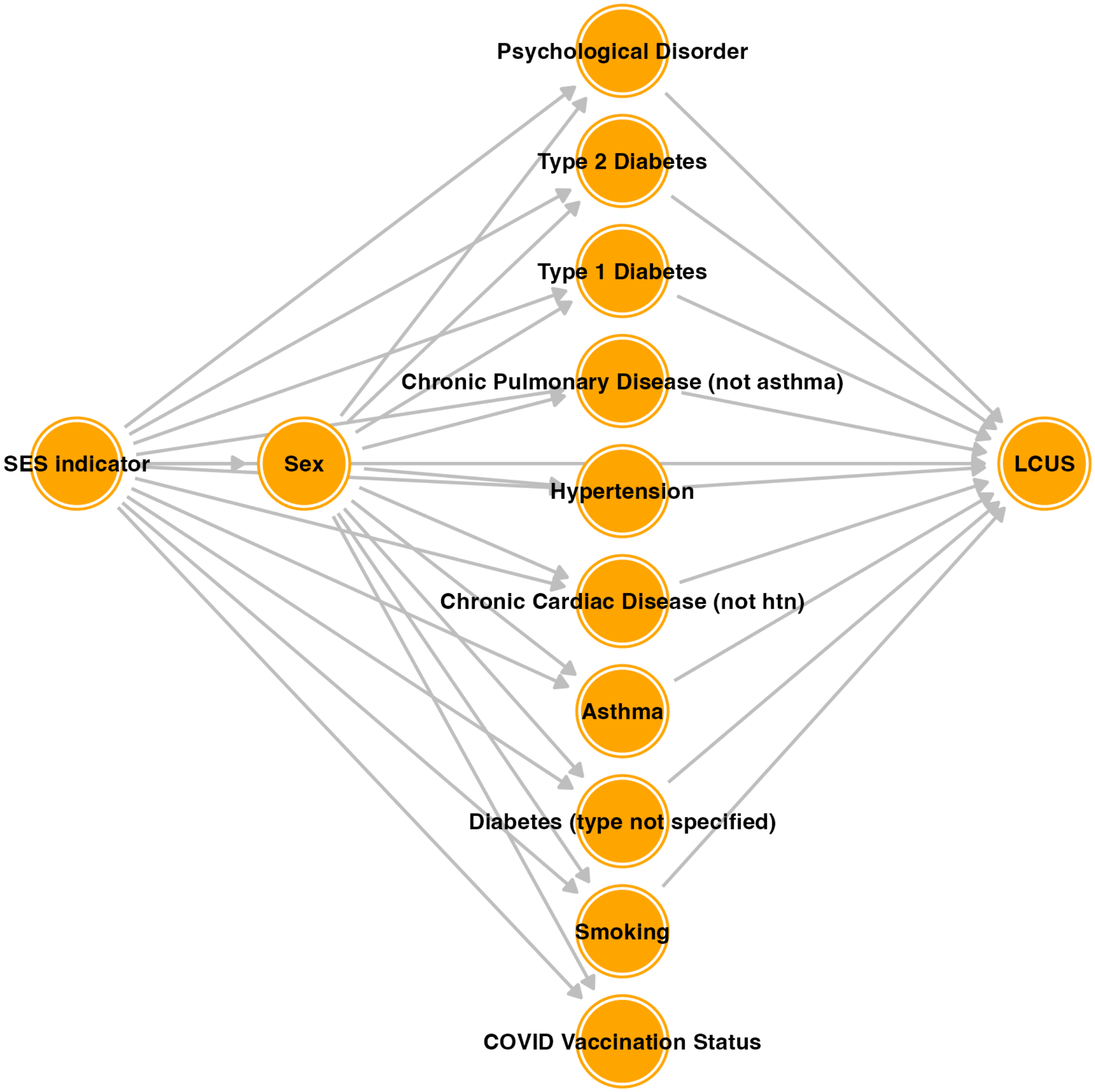

Psychological Disorder
Type 2 Diabetes
Type 1 Diabetes
Chronic Pulmonary Disease (not asthma)
SES indicator
Sex
Hypertension
LCUS
Chronic Cardiac Disease (not htn)
Asthma
Diabetes (type not specified)
Smoking
COVID Vaccination Status

(B)

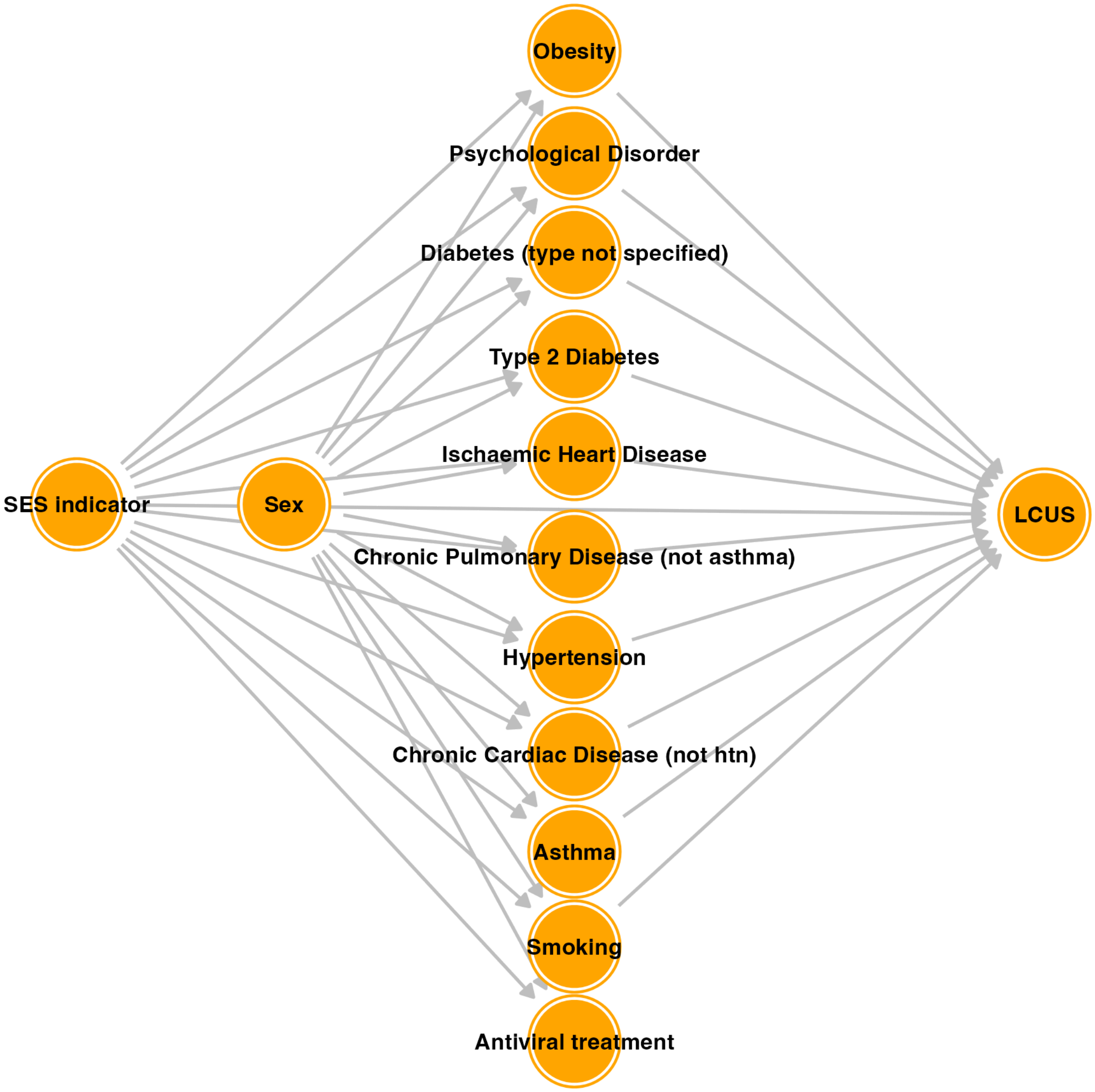
Obesity
Psychological Disorder
Diabetes (type not specified)
Type 2 Diabetes
Ischaemic Heart Disease
SES indicator
Sex
LCUS
Chronic Pulmonary Disease (not asthma)
Hypertension
Chronic Cardiac Disease (not htn)
Asthma
Smoking
Antiviral treatment

**(C)**

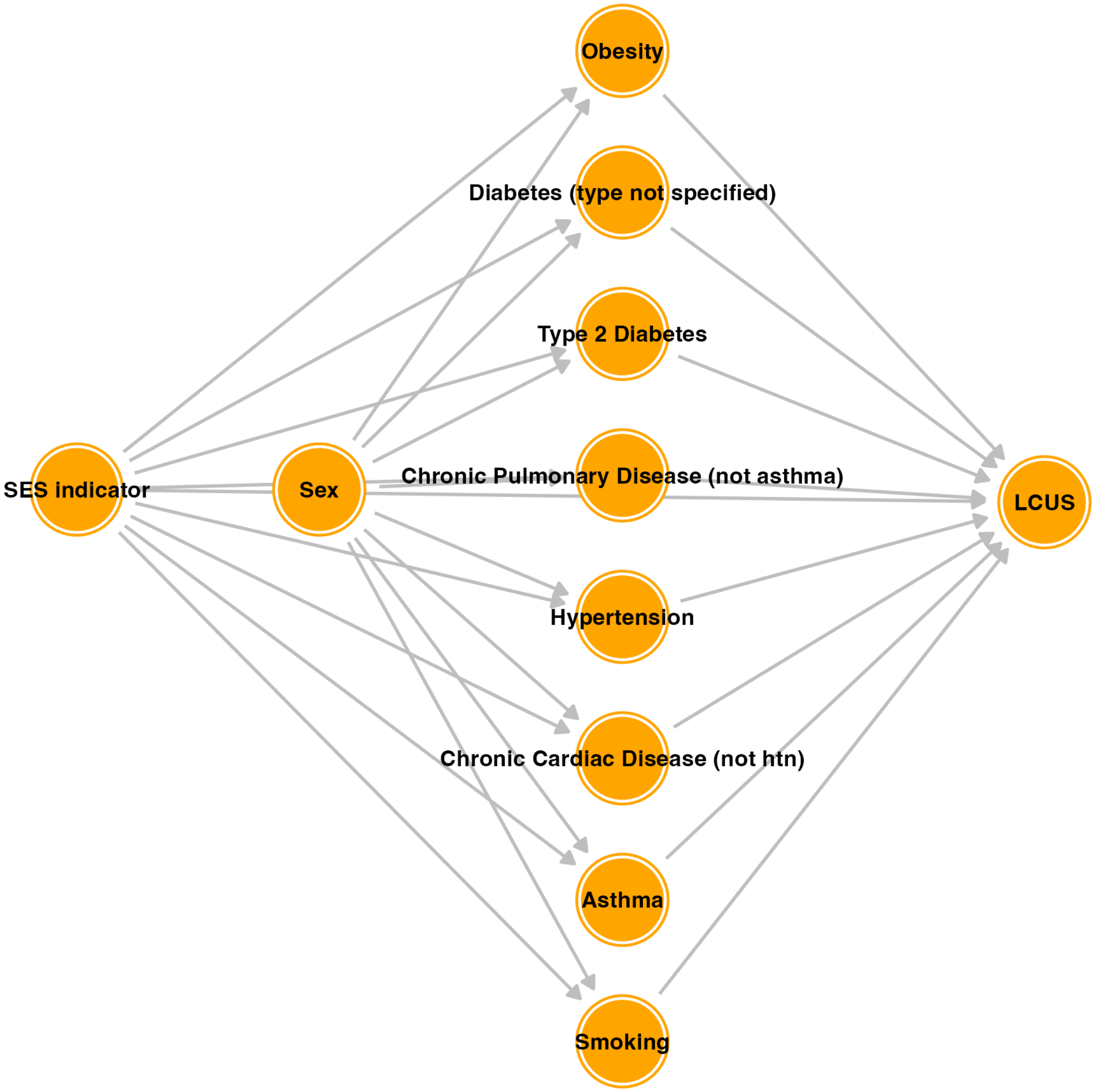


**Supplementary Figure S2. Assumed DAGs for the Norway, UK, and Russia cohorts with Sex (female vs male) as exposure. DAG for Sex in Norway (A), DAG for Sex in the UK (B), DAG for Sex in Russia (C). Note: htn = hypertension, in "Chronic Cardiac Disease (not htn)." LCUS denotes Long COVID utility scores.**

**(A)**

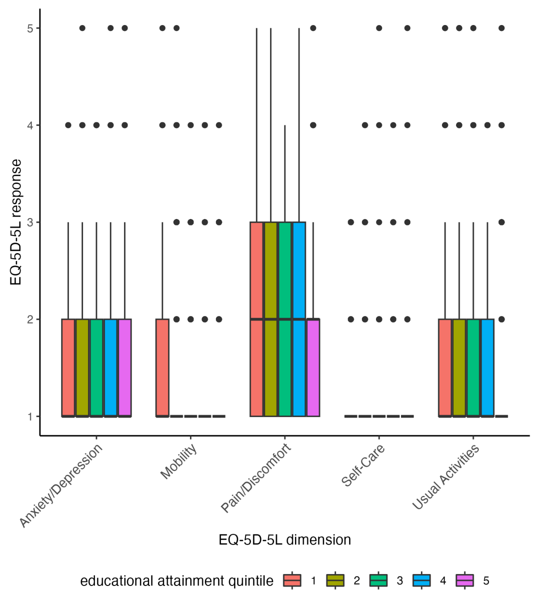


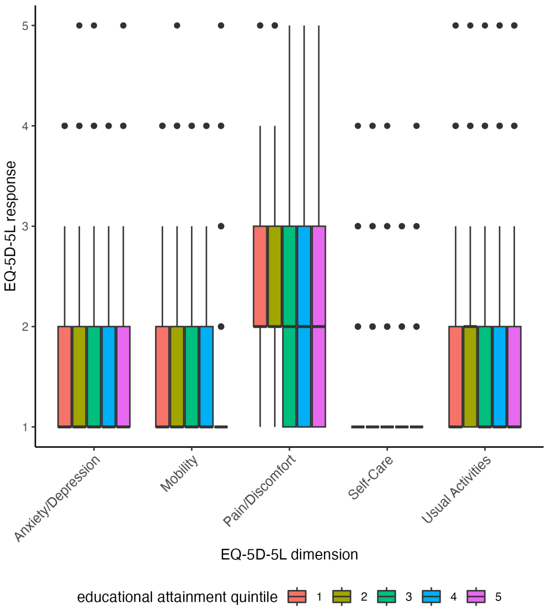


**(i)** **(ii)**

**(B)**

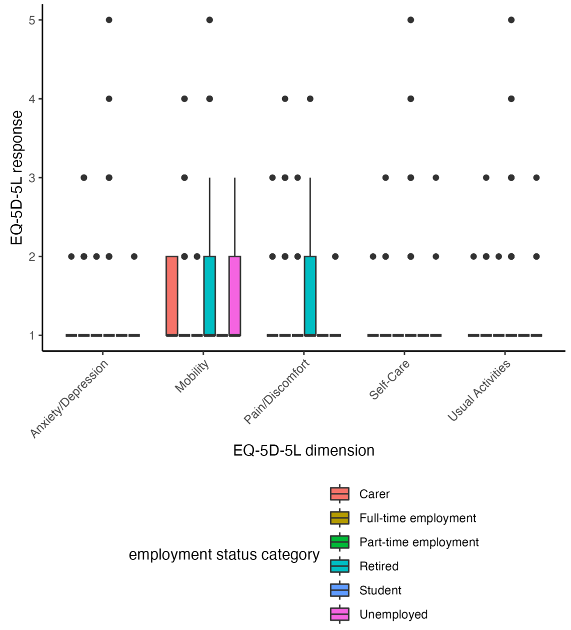


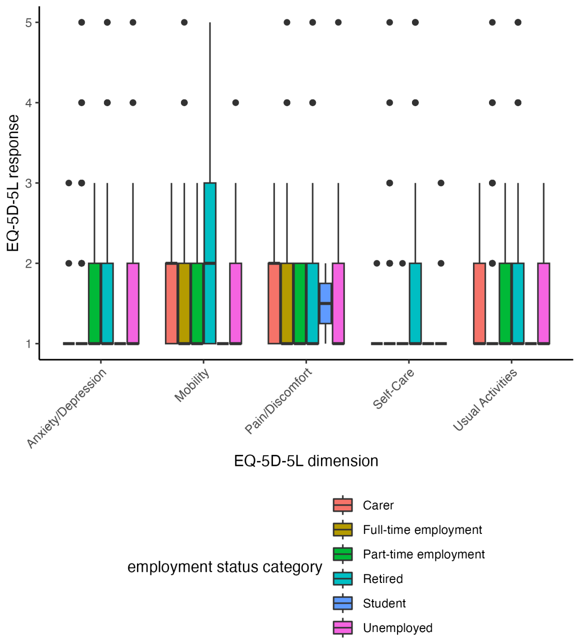


**(i)** **(ii)**

**Supplementary Figure S3. Boxplot comparisons of EQ-5D-5L responses (score of 1-5) by dimension and quintile of educational attainment in Norway, before COVID-19 illness (i) and after COVID-19 illness (ii) (A) and by dimension and employment status category in Russia, before COVID-19 illness (i) and after COVID-19 illness (ii) (B). Bars denote median scores. Points are drawn for scores which lie 1.5*the interquartile range units above the upper quartile.**

**(A)**

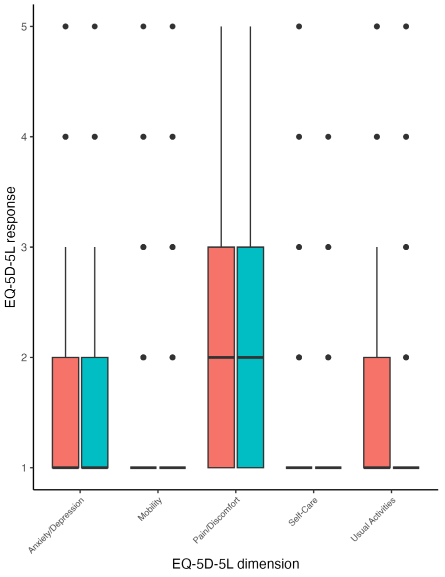


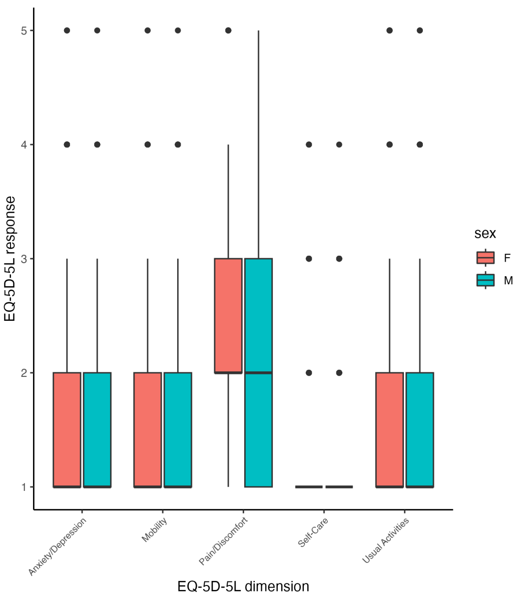


**(i)** **(ii)**

**(B)**

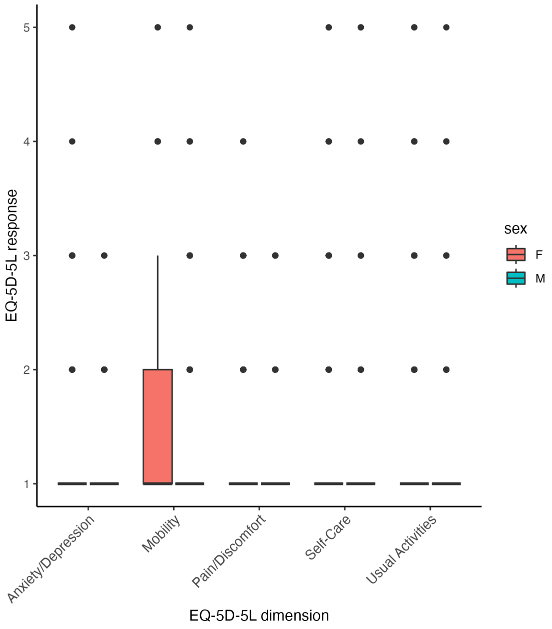


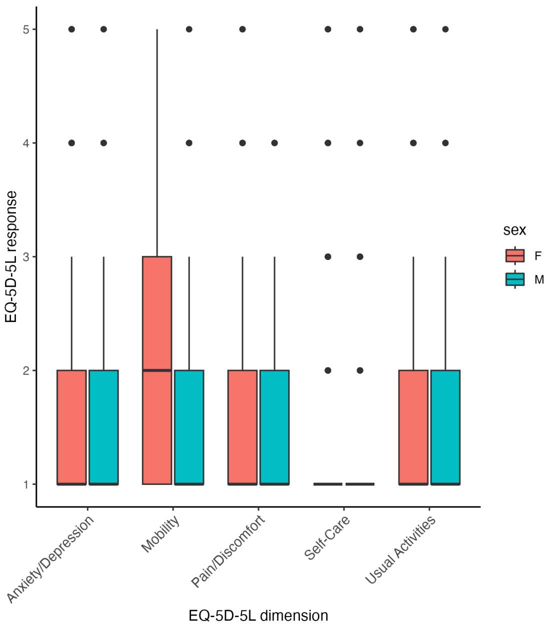


**(i)** **(ii)**

**Supplementary Figure S4. Boxplot comparisons of EQ-5D-5L responses by dimension and sex (Female (F), Male (M), and Unknown (U)) in Norway, before COVID-19 illness (i) and after COVID-19 illness (ii) (A), the UK (i and ii) (B), and Russia (i and ii) (C). Points are drawn for scores which lie 1.5*the interquartile range units above the upper quartile.**

**(A)**

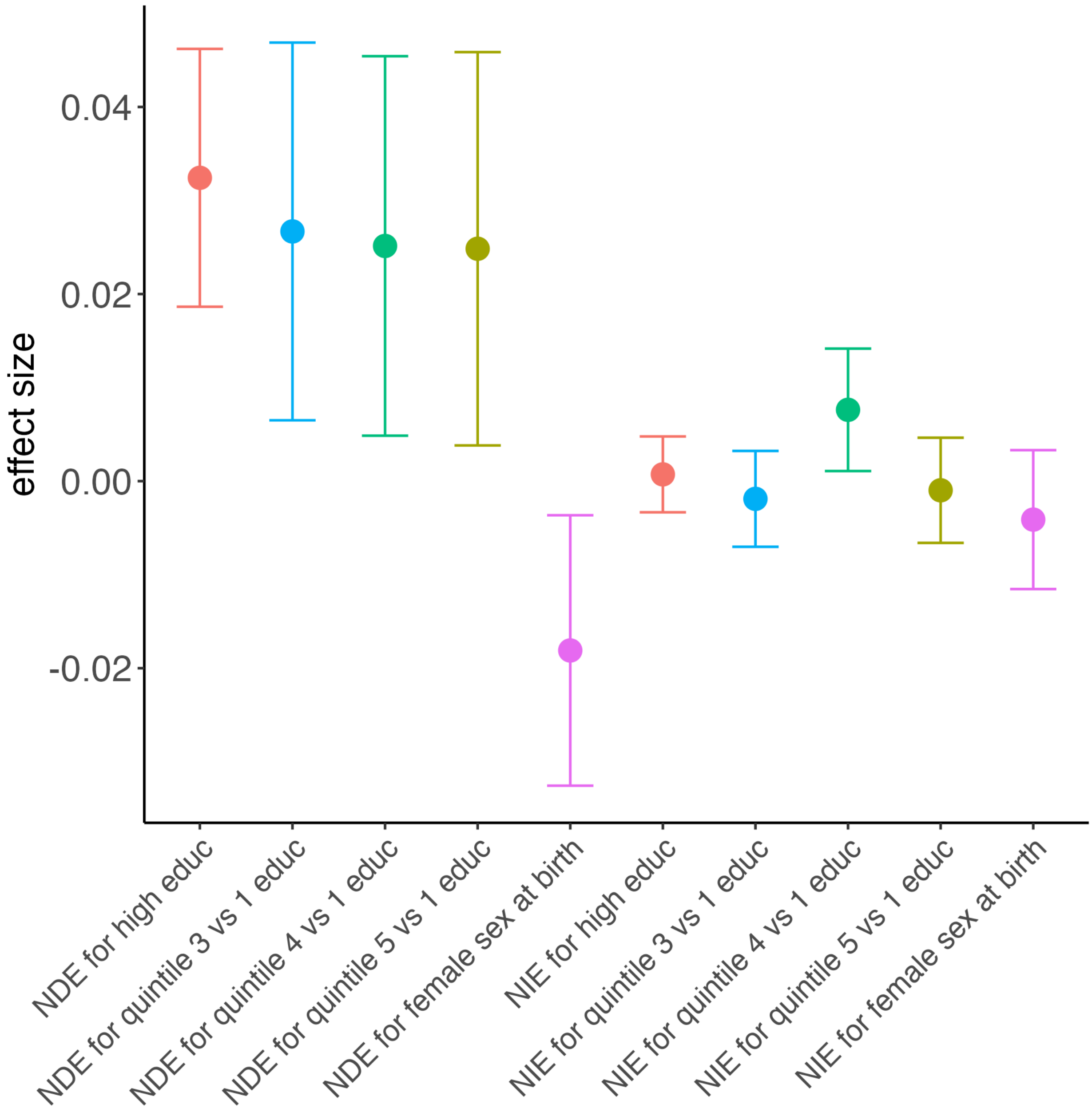

effect size
0.04
0.02
0.00
-0.02
NDE for high educ
NDE for quintile 3 vs 1 educ
NDE for quintile 4 vs 1 educ
NDE for quintile 5 vs 1 educ
NDE for female sex at birth
NIE for high educ
NIE for quintile 3 vs 1 educ
NIE for quintile 4 vs 1 educ
NIE for quintile 5 vs 1 educ
NIE for female sex at birth

(B)

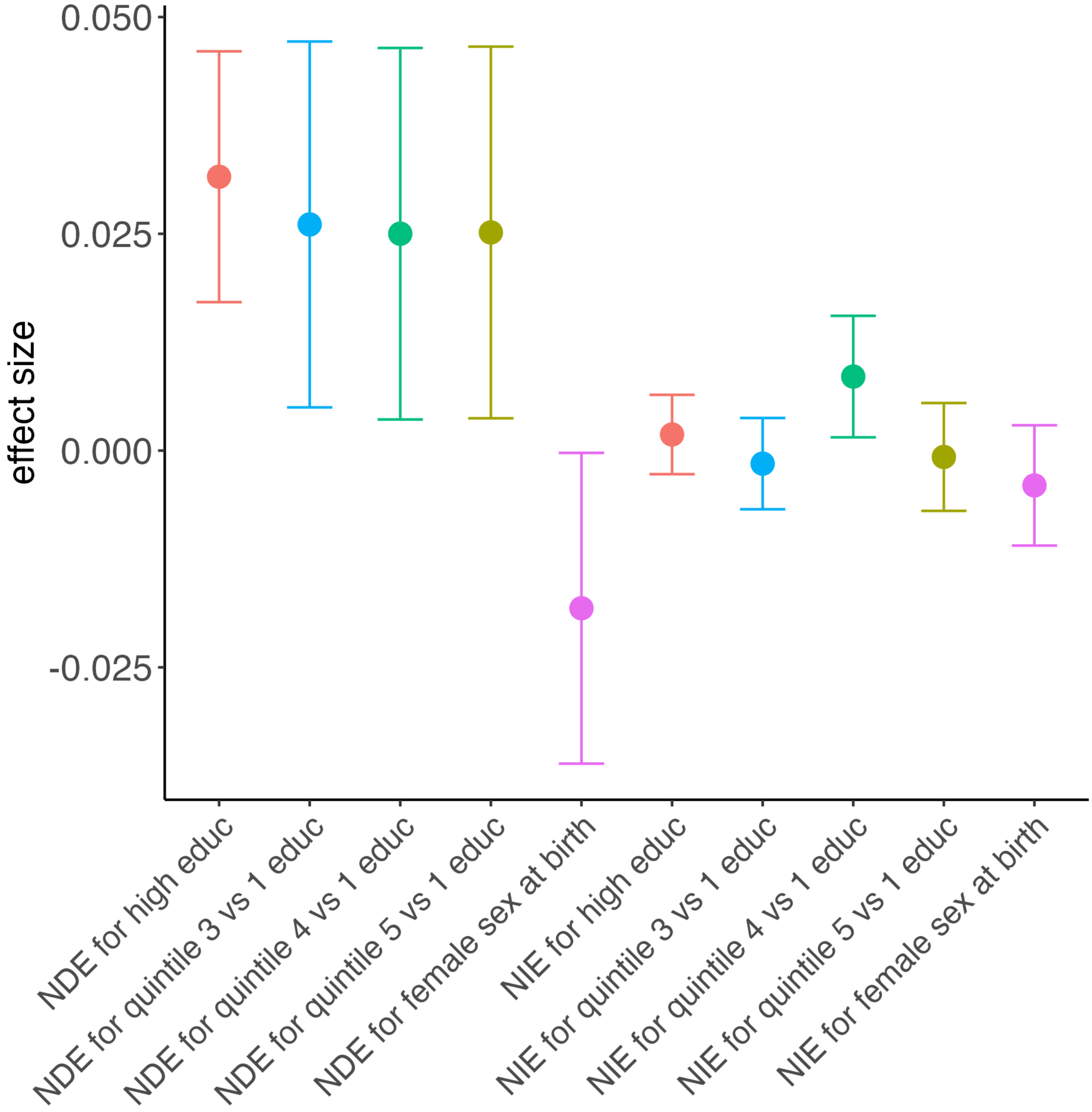
0.050
0.025
0.000
-0.025
effect size
NDE for high educ
NDE for quintile 3 vs 1 educ
NDE for quintile 4 vs 1 educ
NDE for quintile 5 vs 1 educ
NDE for female sex at birth
NIE for high educ
NIE for quintile 3 vs 1 educ
NIE for quintile 4 vs 1 educ
NIE for quintile 5 vs 1 educ
NIE for female sex at birth

**(C)**

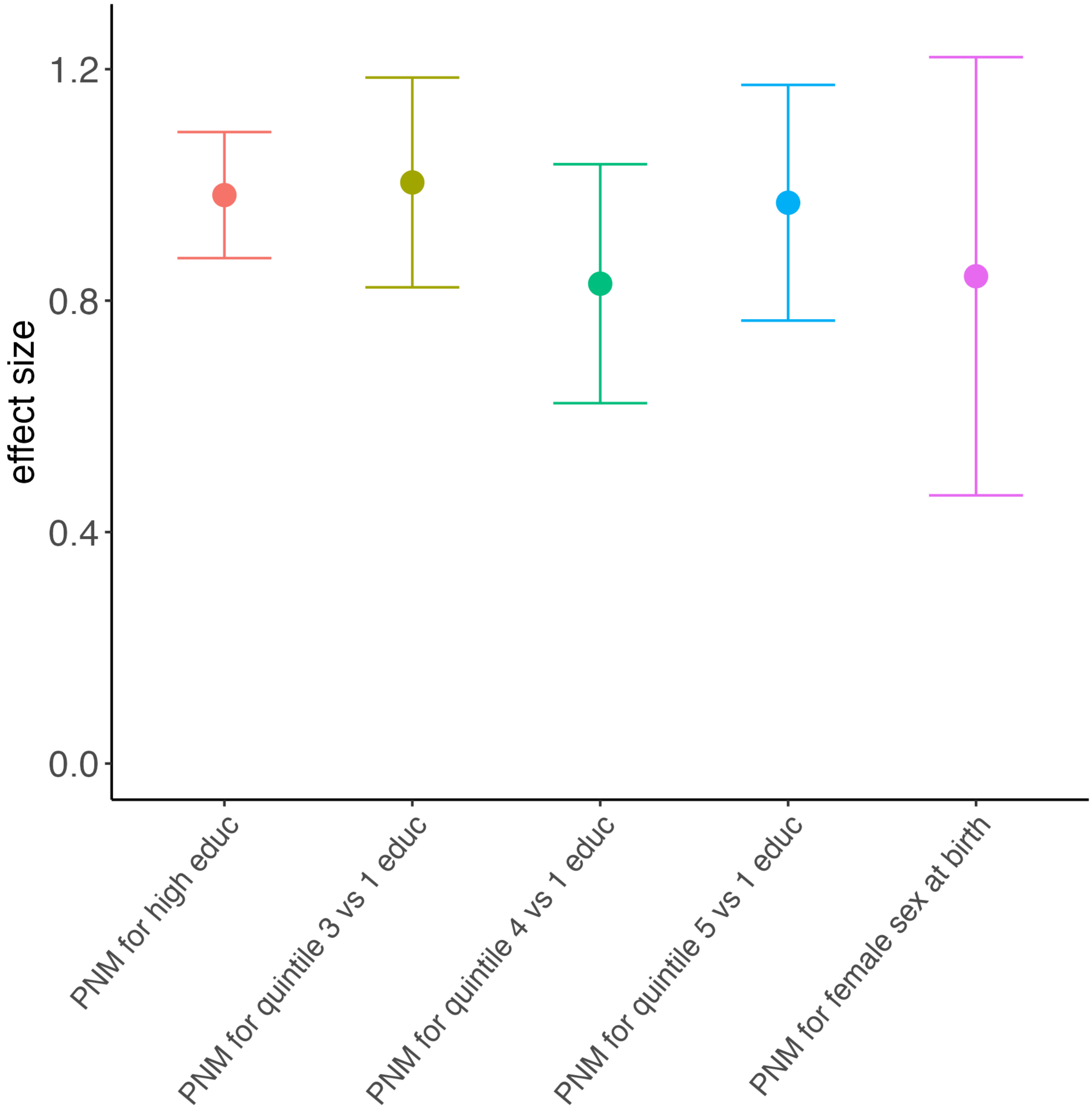

1.2
0.8
0.4
0.0
effect size
PNM for high educ
PNM for quintile 3 vs 1 educ
PNM for quintile 4 vs 1 educ
PNM for quintile 5 vs 1 educ
PNM for female sex at birth

**(D)**

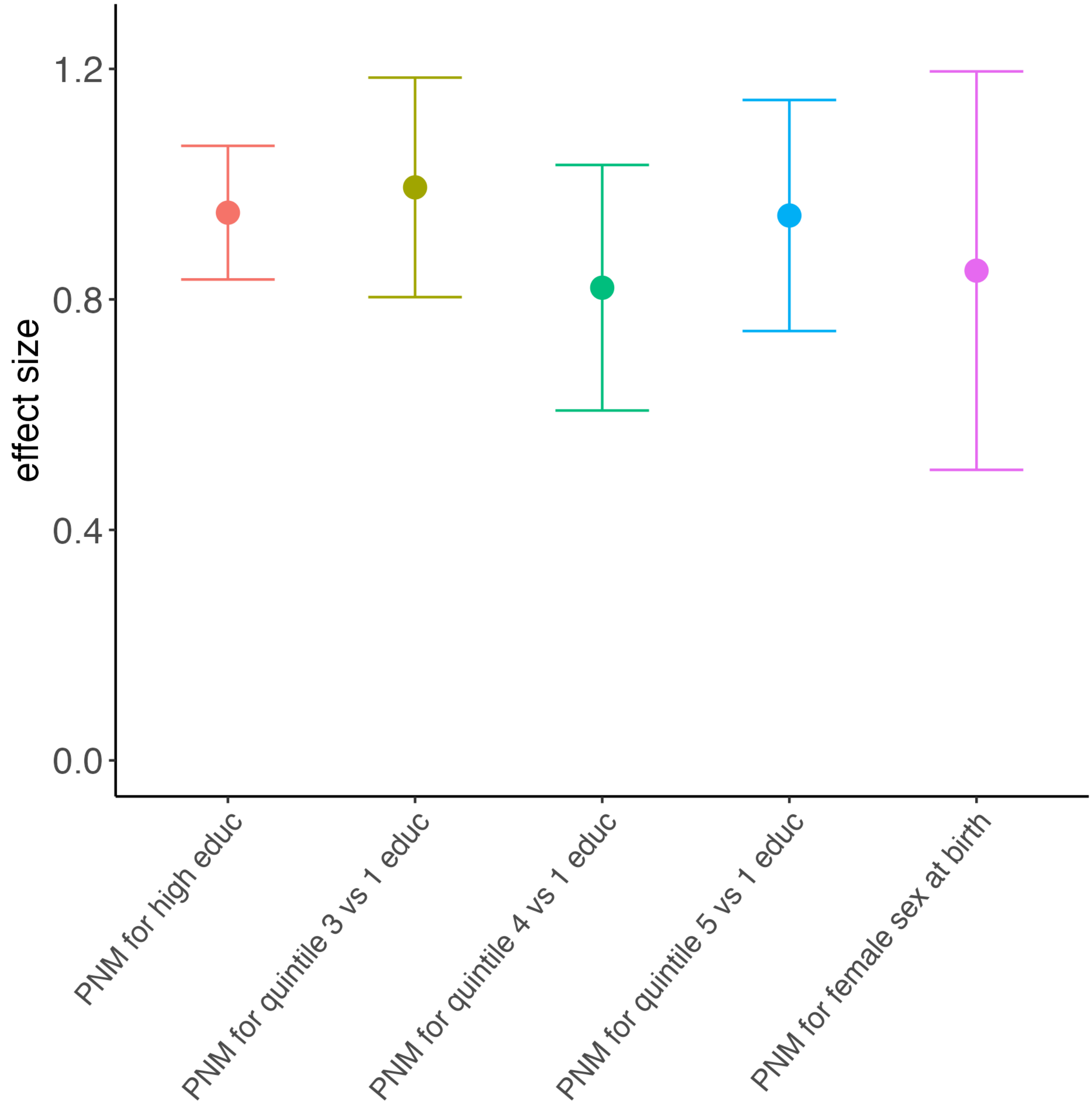


**Supplementary Figure S5. All output for Norway mediation analyses. Estimated NDEs and NIEs (point estimate and 95% confidence interval) in the main analysis (A) and in the sensitivity analysis (B). Colors denote the contrast of interest (pink: high vs low educational attainment; blue: quintile 3 vs 1 of educational attainment; green: quintile 4 vs 1 of educational attainment; khaki: quintile 5 vs 1 of educational attainment; purple: female vs male sex). Estimated proportions non-mediated (point estimate and 95% confidence interval) in the main analysis (C) and in the sensitivity analysis (D).**

**(A)**

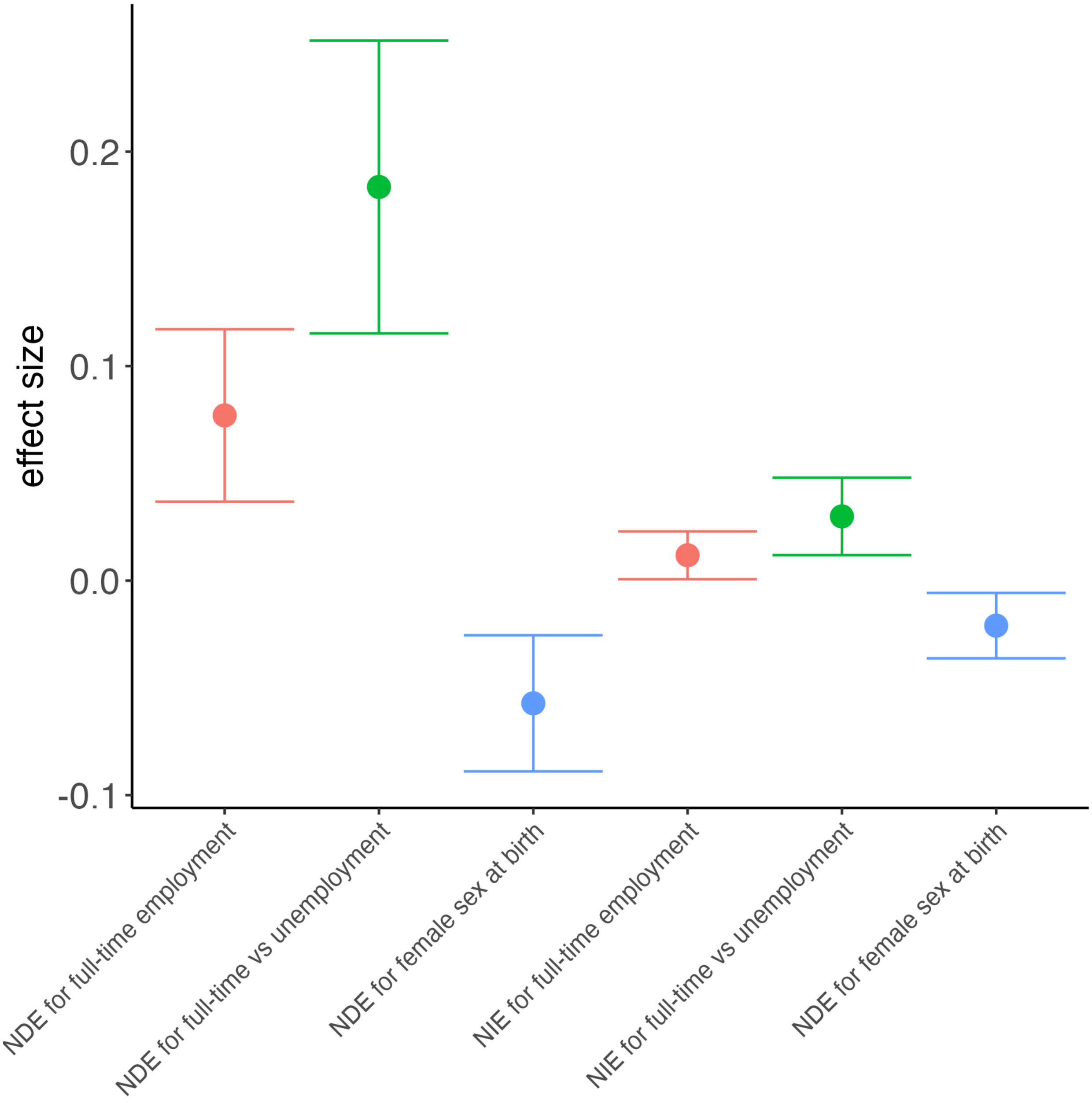

effect size
0.2
0.1
0.0
-0.1
NDE for full-time employment
NDE for full-time vs unemployment
NDE for female sex at birth
NIE for full-time employment
NIE for full-time vs unemployment
NDE for female sex at birth

**(B)**

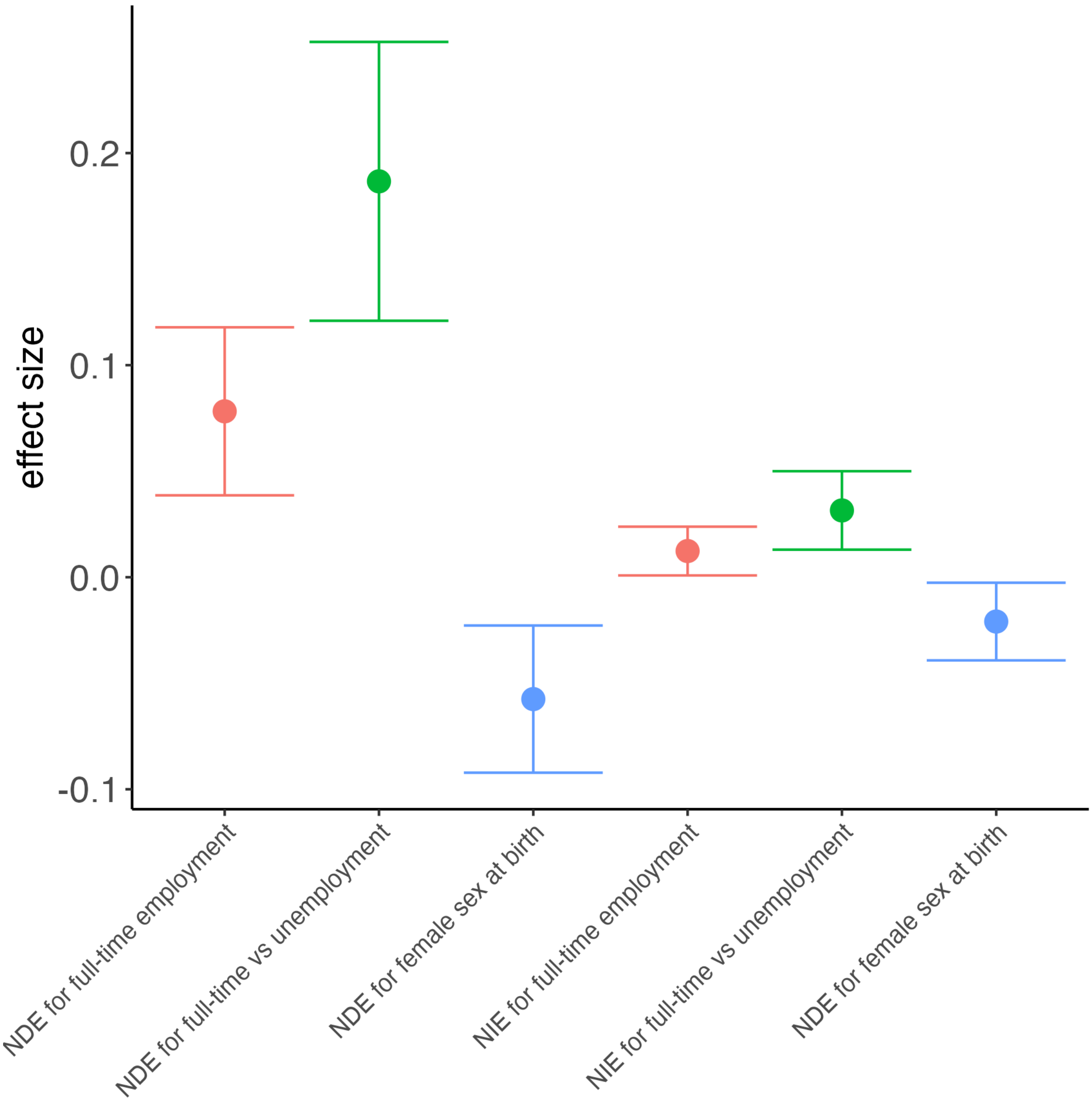

effect size
0.2
0.1
0.0
-0.1
NDE for full-time employment
NDE for full-time vs unemployment
NDE for female sex at birth
NIE for full-time employment
NIE for full-time vs unemployment
NDE for female sex at birth

**(C)**

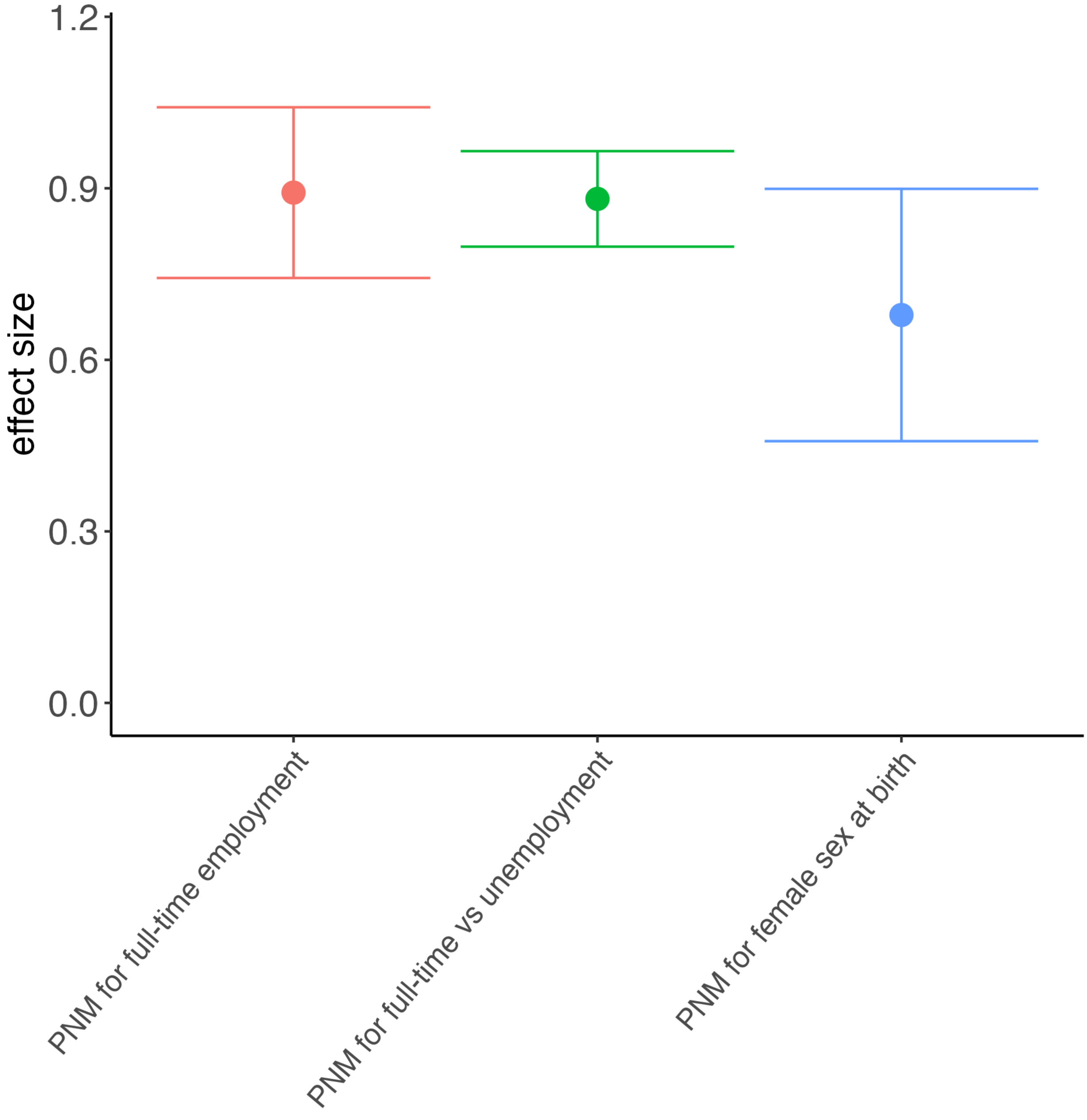
1.2
0.9
0.6
0.3
0.0
effect size
PNM for full-time employment
PNM for full-time vs unemployment
PNM for female sex at birth

**(D)**

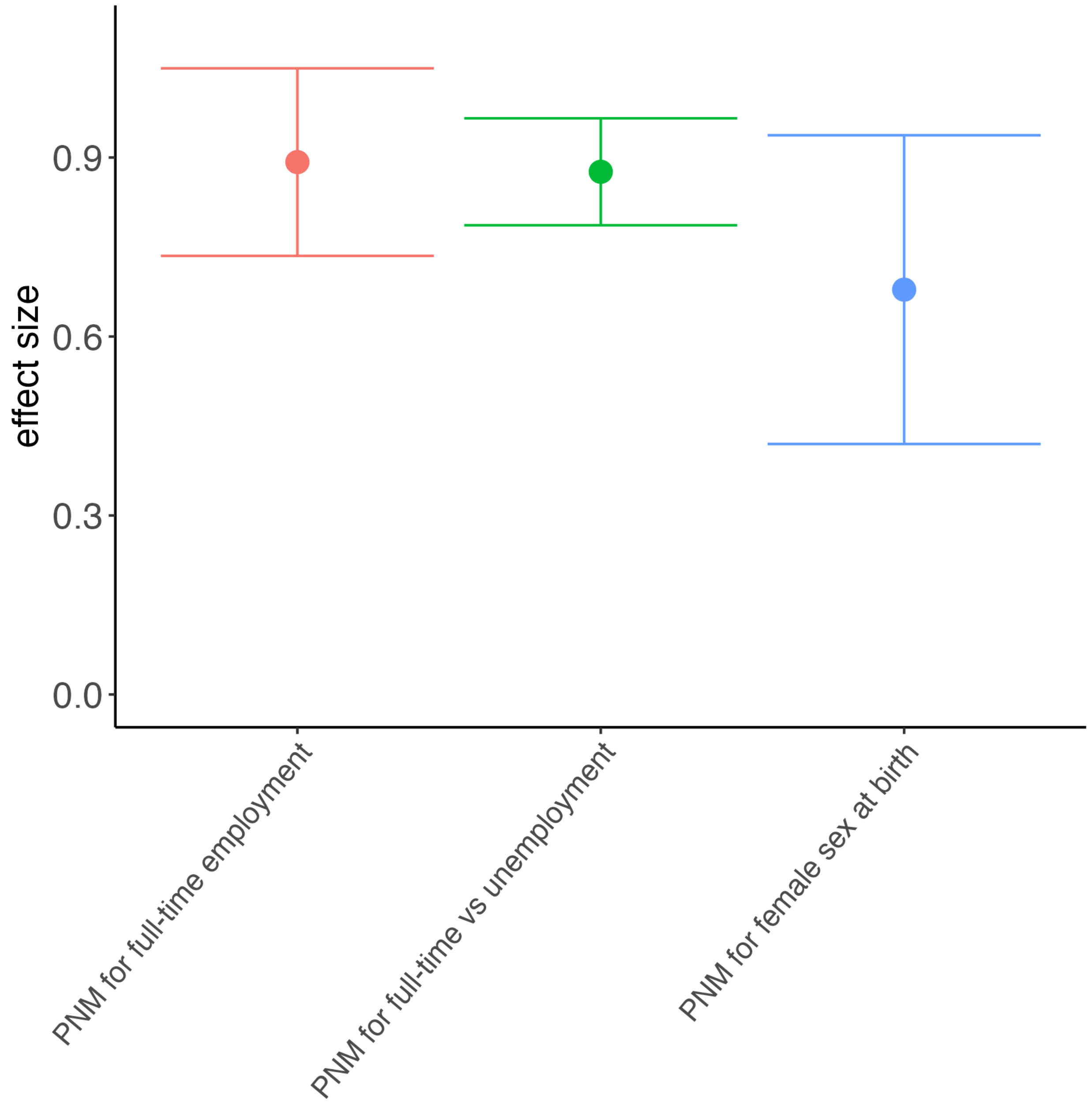


**Supplementary Figure S6. All output for UK mediation analyses. Estimated NDEs and NIEs (point estimate and 95% confidence interval) in the main analysis (A) and in the sensitivity analysis (B). Colors denote the contrast of interest (pink: full-time employment vs all other employment status categories; green: full-time employment vs unemployment; blue: female vs male sex). Estimated proportions non-mediated (point estimate and 95% confidence interval) in the main analysis (C) and in the sensitivity analysis (D).**

**(A)**

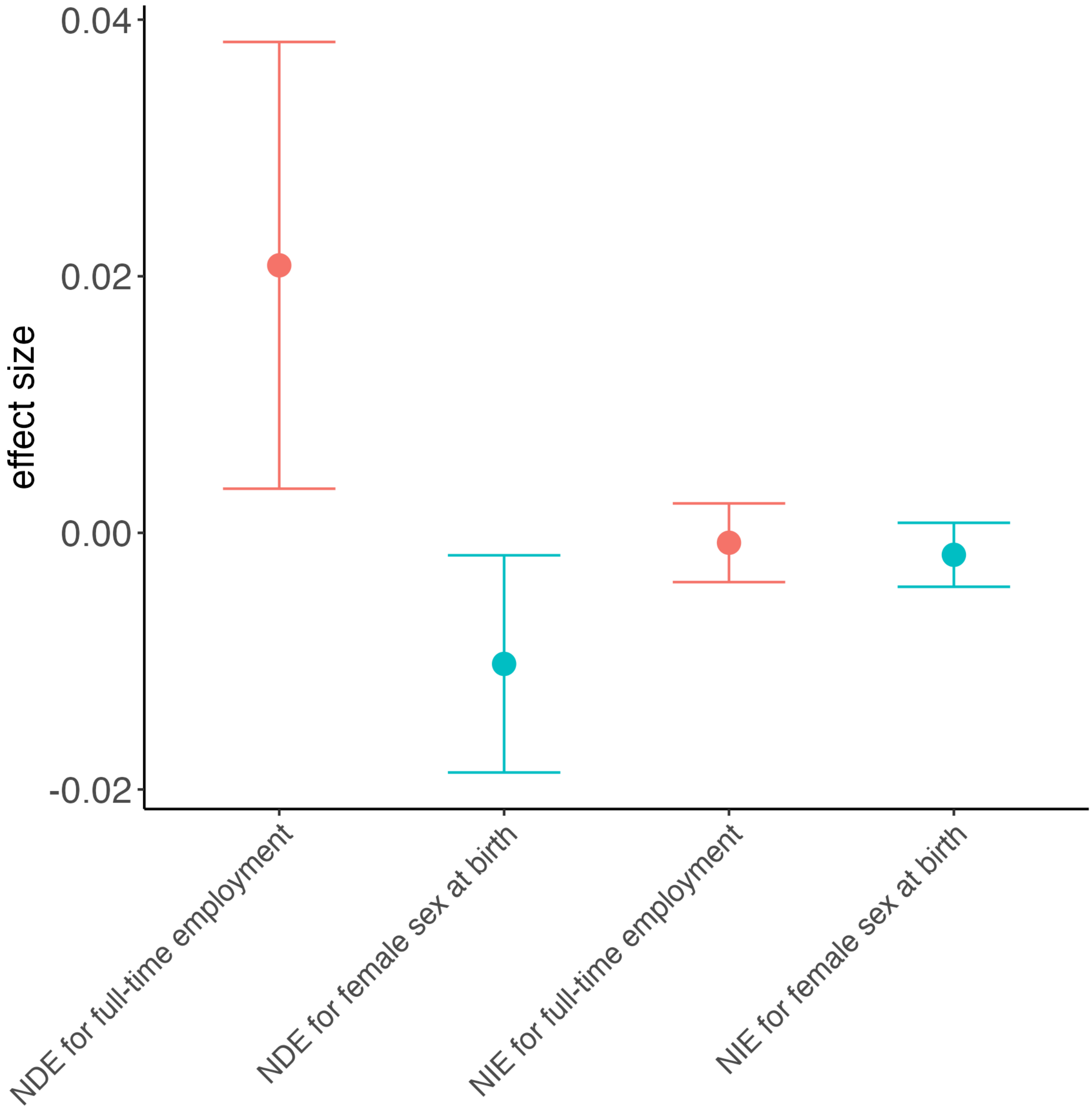

0.04
0.02
0.00
-0.02
effect size
NDE for full-time employment
NDE for female sex at birth
NIE for full-time employment
NIE for female sex at birth

**(B)**

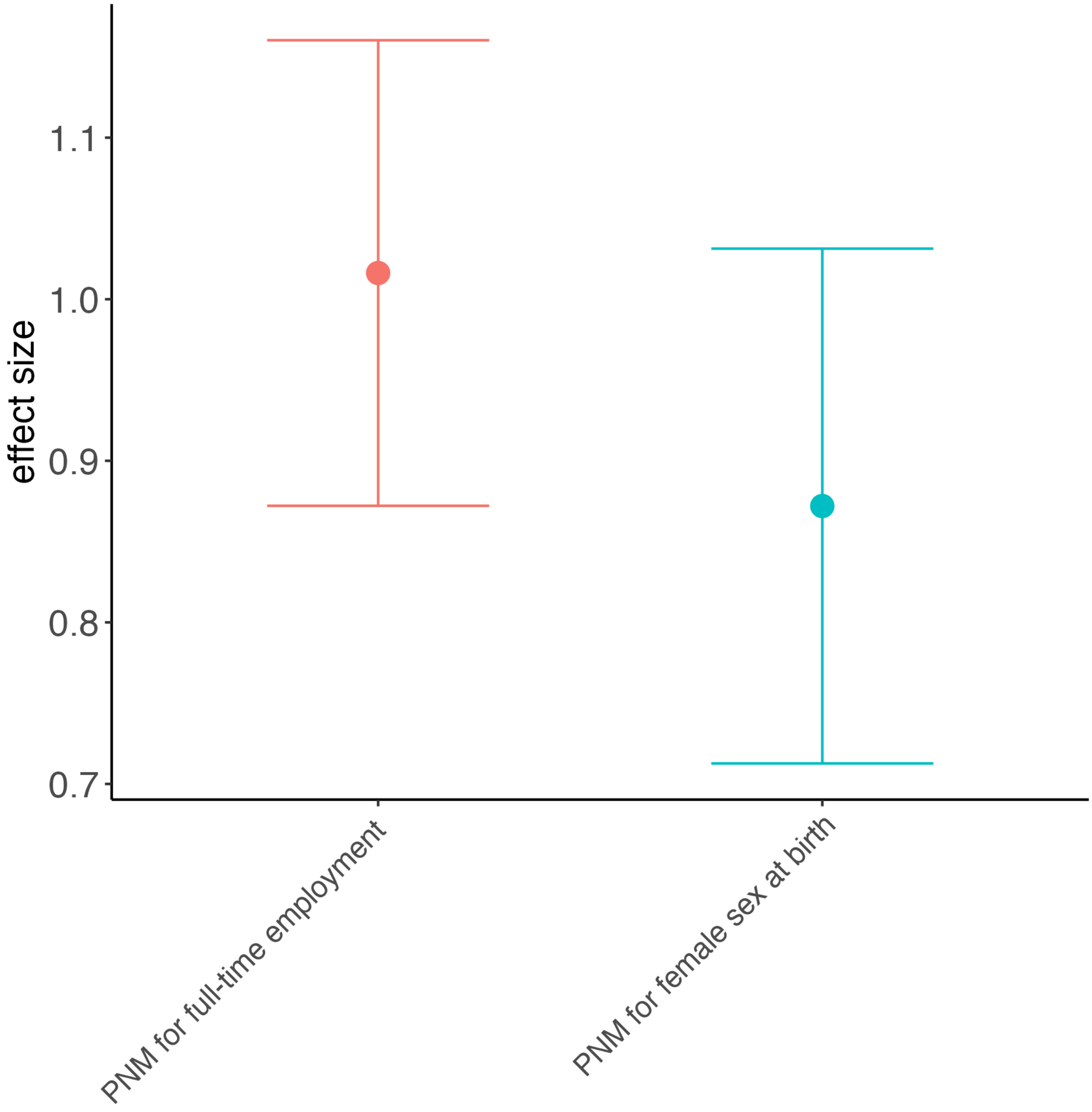


**Supplementary Figure S7. All output for the Russia mediation analyses. Estimated NDEs and NIEs (point estimate and 95% confidence interval) (A). Colors denote the contrast of interest (pink: full-time employment vs all other employment status categories; cyan: female vs male sex). Estimate proportions non-mediated (B).**

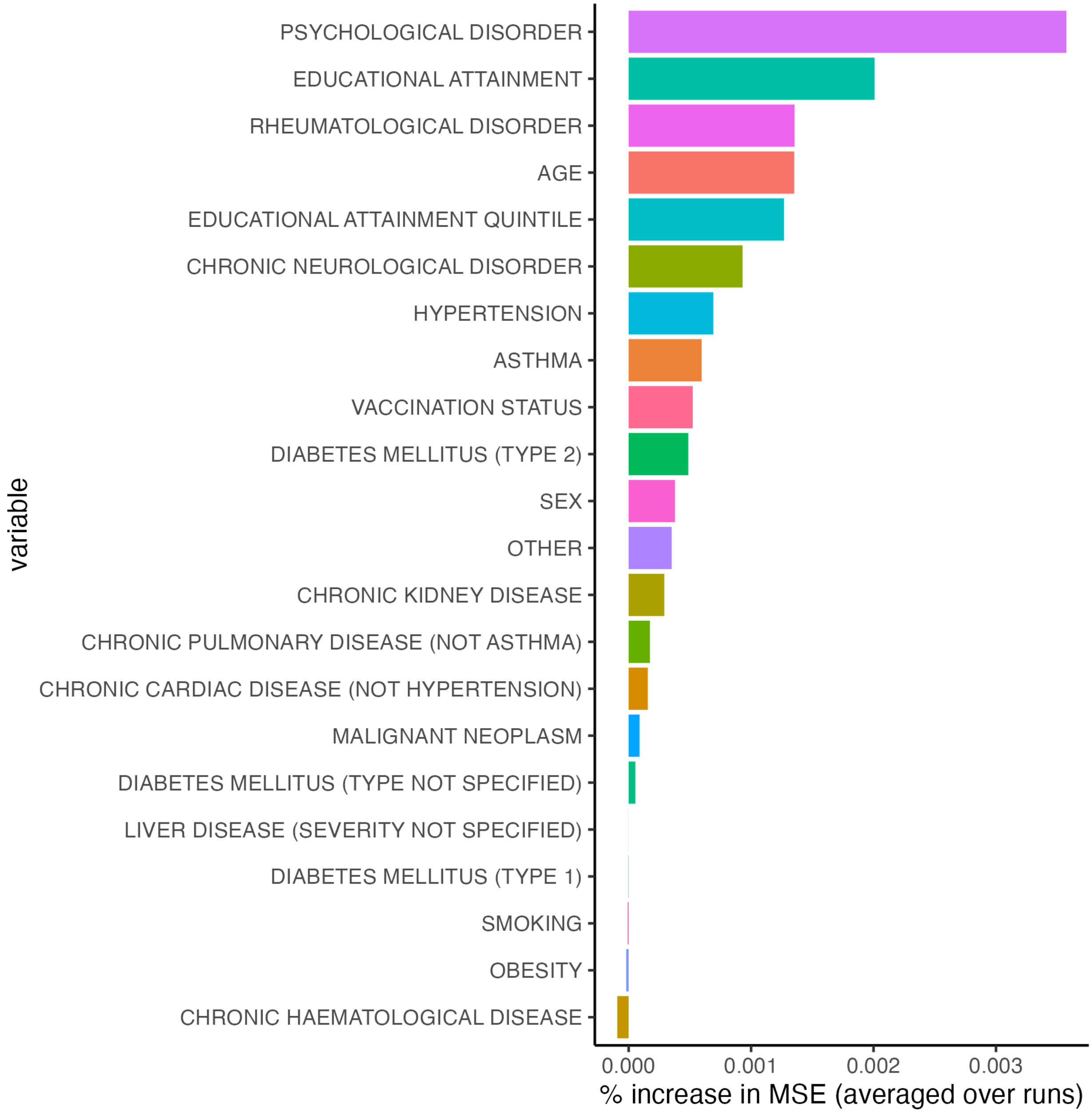


**Supplementary Figure S8A. Estimated variable importance measures, i.e. % increase in mean squared error or MSE, from individual random forest implementation (RF #1) for Norway (sex-based population correction sensitivity analysis).**

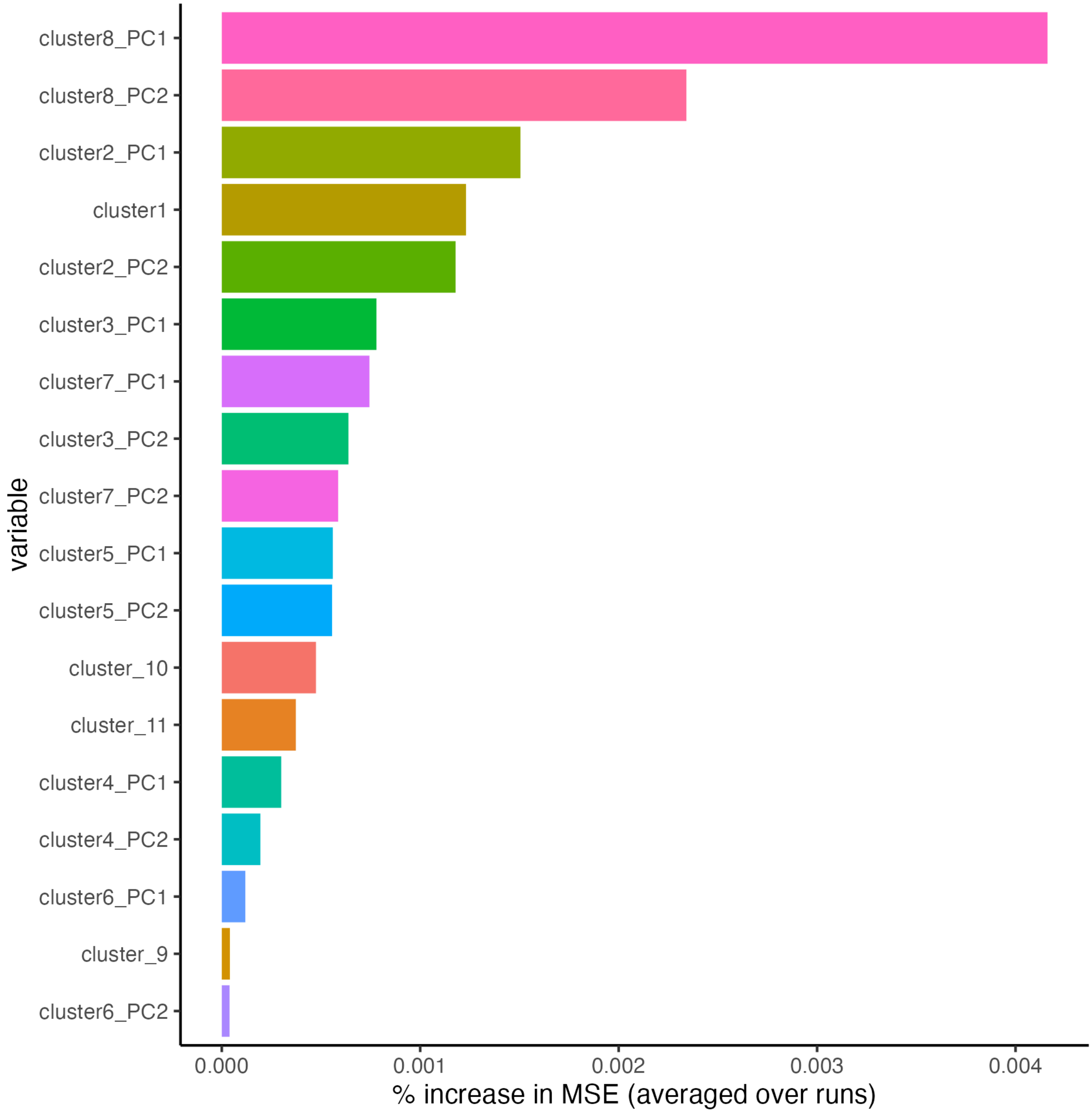


**Supplementary Figure 8B. Estimated variable importance measures, i.e. % increase in mean squared error or MSE, from pre-grouped random forest implementation (RF #2) for Norway. Rows indicate cluster names (a full list of variables belonging to each cluster can be found in Supplementary Table S3) and corresponding principal components, if the cluster consists of multiple variables. PC1 denotes principal component 1 and PC2 denotes principal component 2 (sex-based population correction sensitivity analysis).**

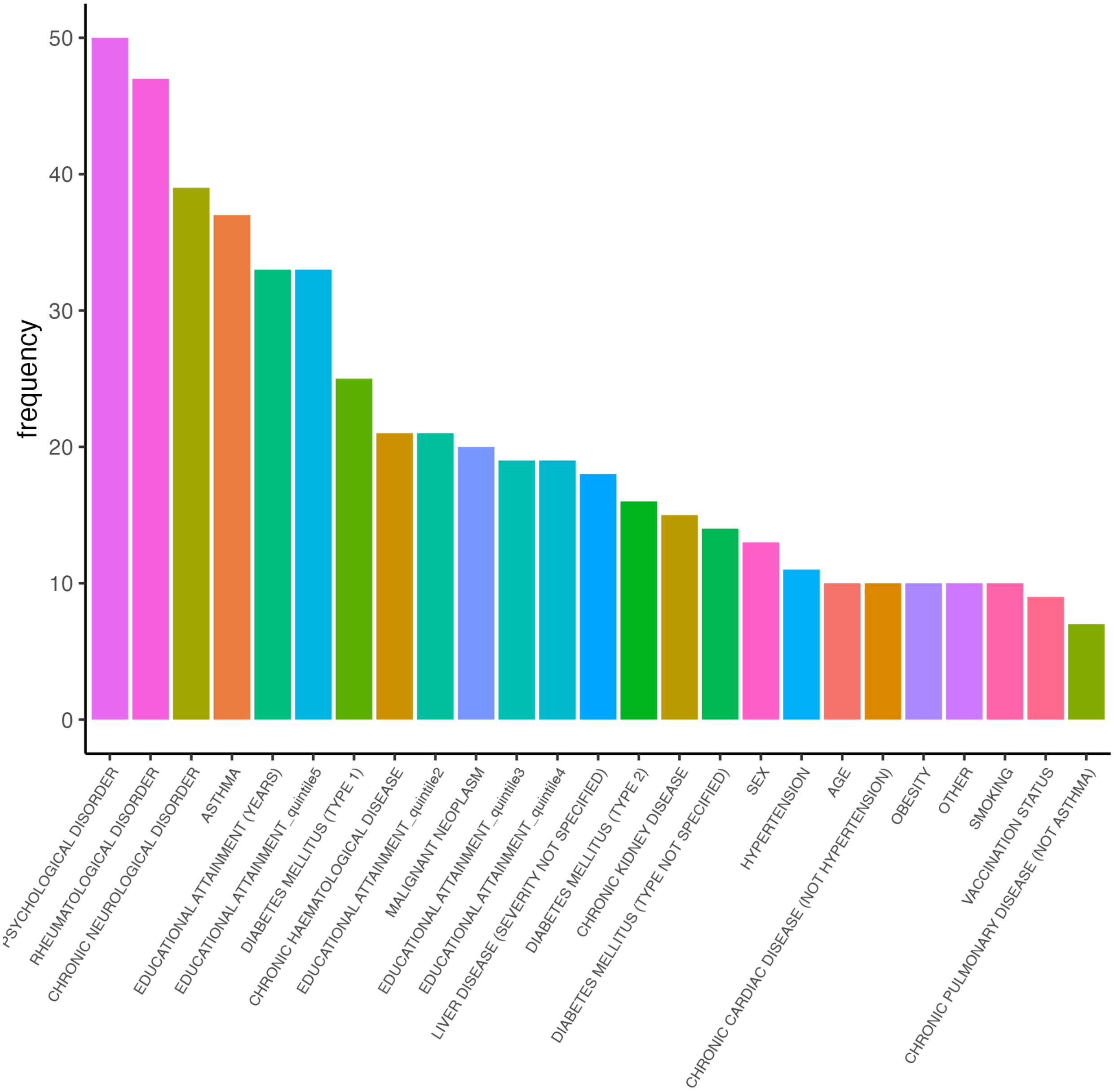

frequency
50
40
30
20
10
0
variable
PSYCHOLOGICAL DISORDER
RHEUMATOLOGICAL DISORDER
CHRONIC NEUROLOGICAL DISORDER
ASTHMA
EDUCATIONAL ATTAINMENT (YEARS)
EDUCATIONAL ATTAINMENT_quintile5
DIABETES MELLITUS (TYPE 1)
CHRONIC HAEMATOLOGICAL DISEASE
EDUCATIONAL ATTAINMENT_quintile2
MALIGNANT NEOPLASM
EDUCATIONAL ATTAINMENT_quintile3
EDUCATIONAL ATTAINMENT_quintile4
LIVER DISEASE (SEVERITY NOT SPECIFIED)
DIABETES MELLITUS (TYPE 2)
CHRONIC KIDNEY DISEASE
DIABETES MELLITUS (TYPE NOT SPECIFIED)
SEX
HYPERTENSION
AGE
CHRONIC CARDIAC DISEASE (NOT HYPERTENSION)
OBESITY
OTHER
SMOKING
VACCINATION STATUS
CHRONIC PULMONARY DISEASE (NOT ASTHMA)

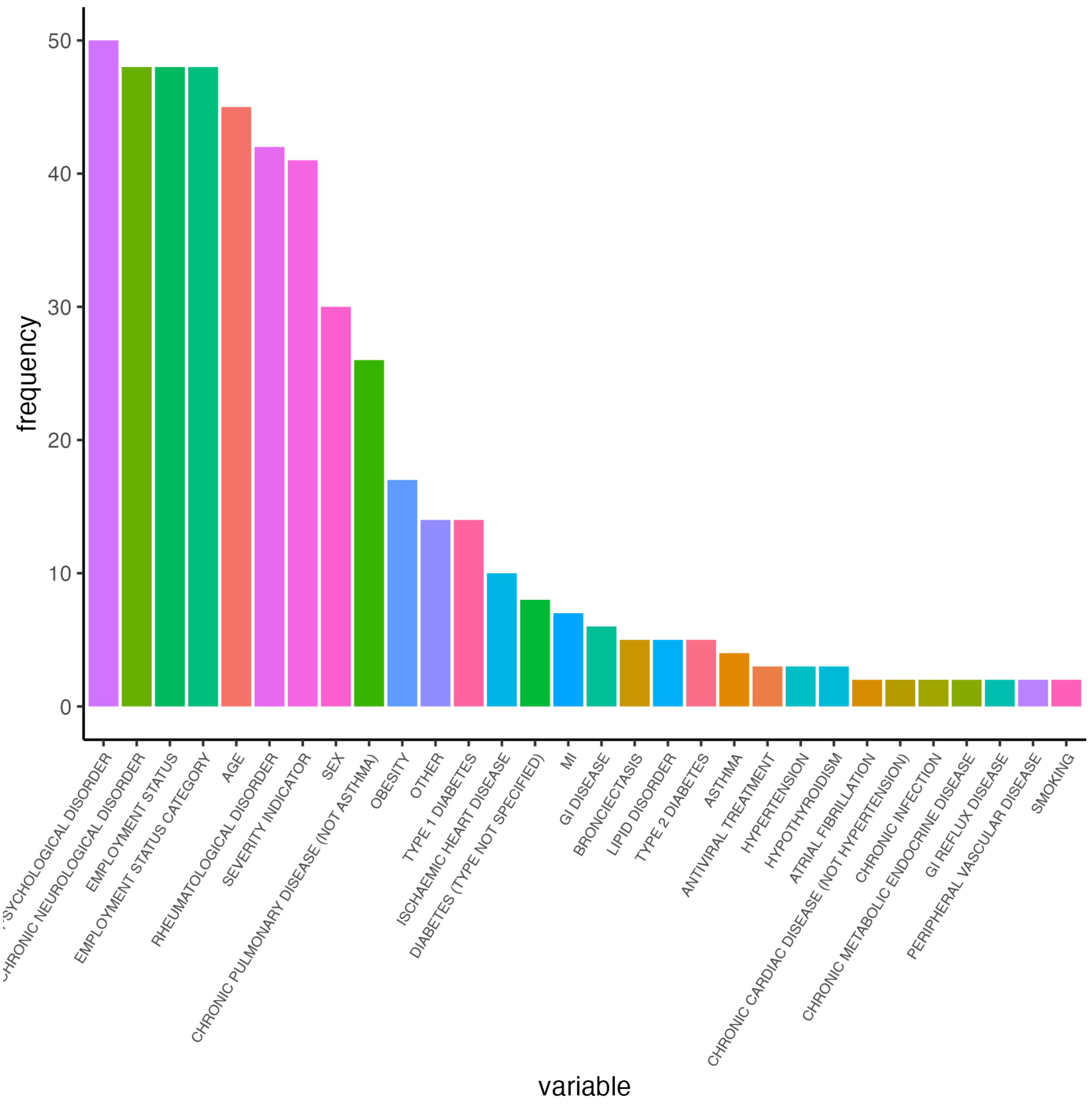


**Supplementary Figure S8C. Number of times (frequency) each variable appears in clusters selected for each CoV-VSURF run (RF #3) for Norway (sex-based population correction sensitivity analysis).**

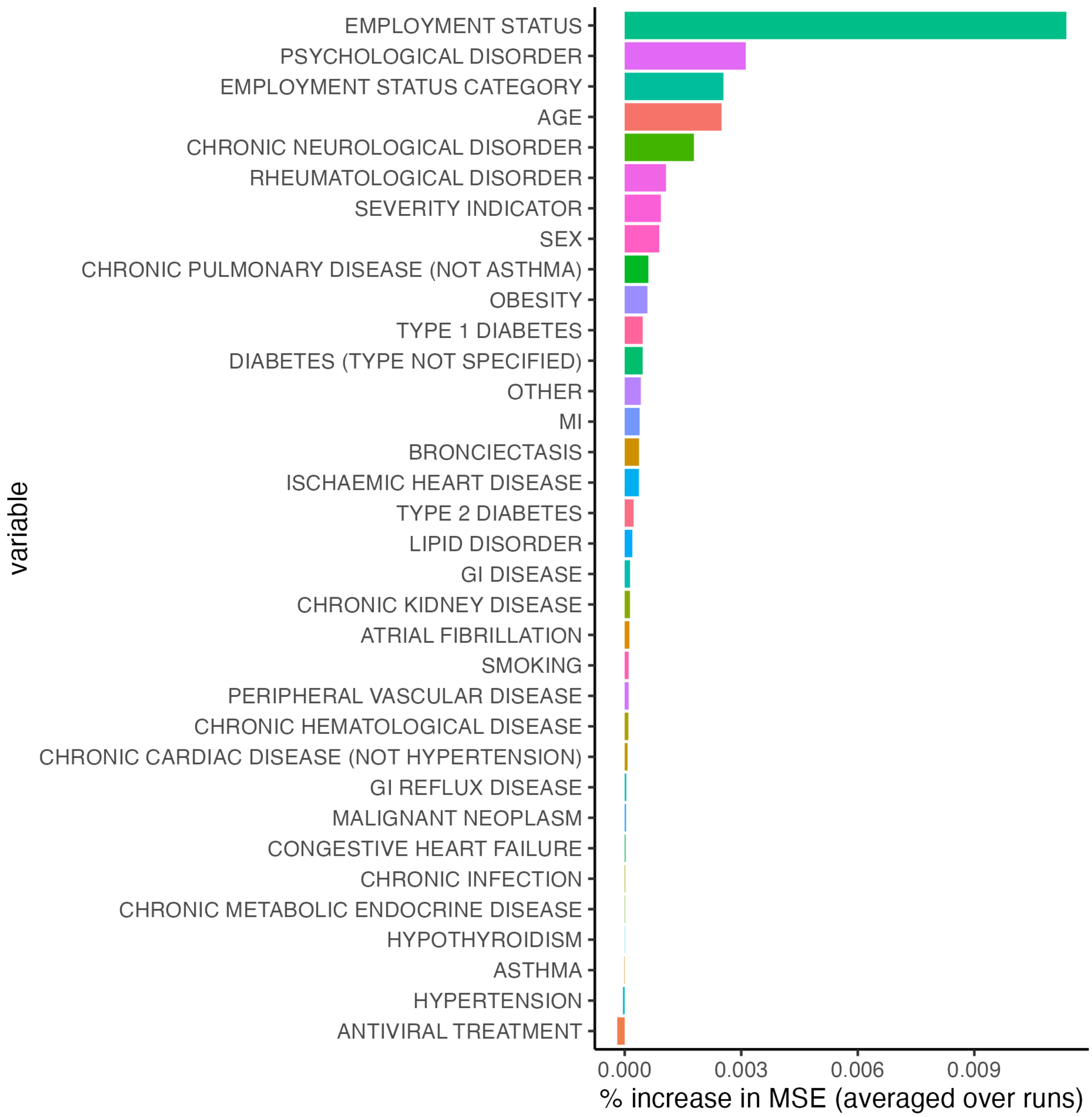


**Supplementary Figure S9A. Estimated variable importance measures, i.e. % increase in mean squared error or MSE, from individual random forest implementation (RF #1) for the UK (sex-based population correction sensitivity analysis).**

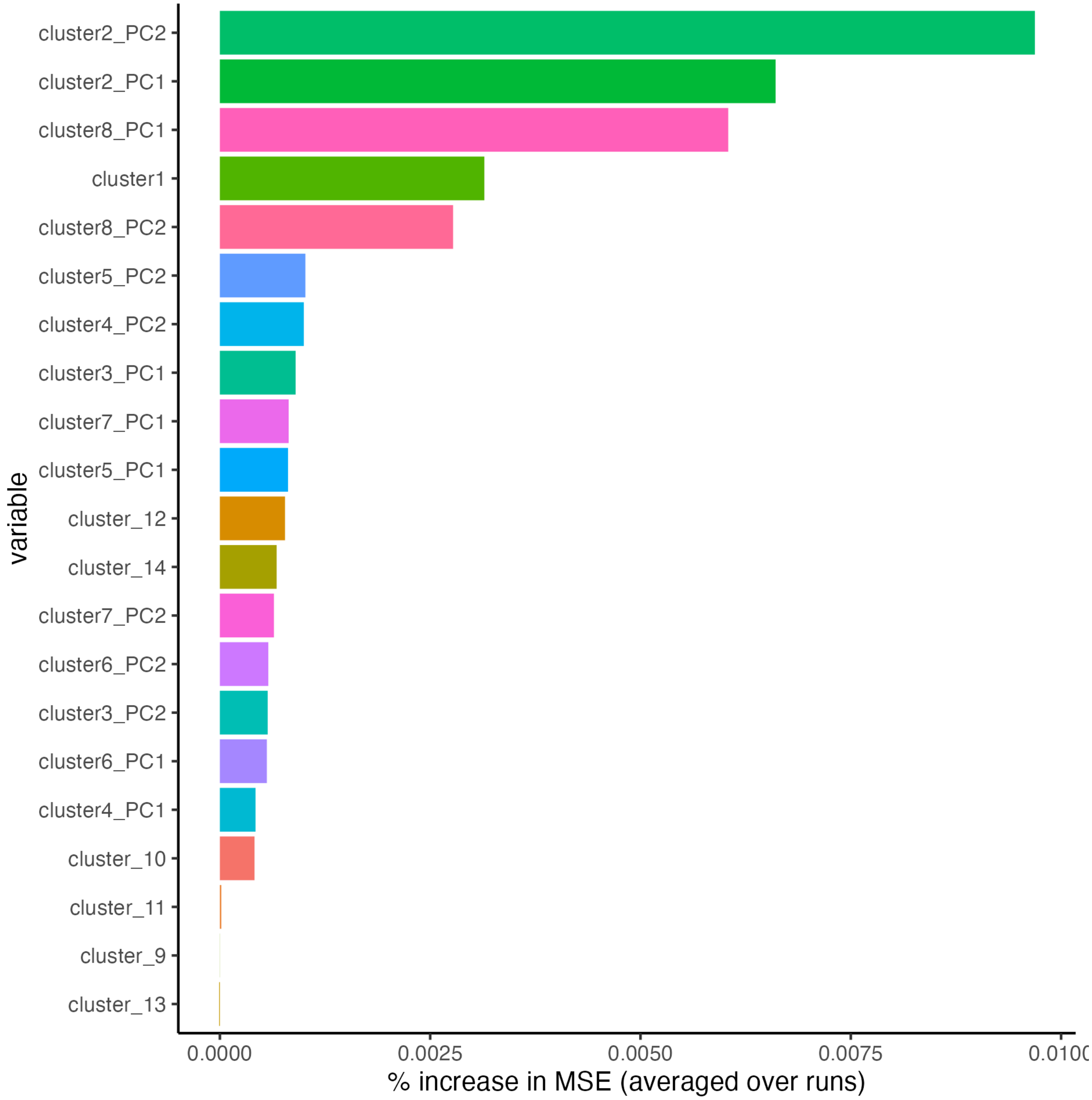


**Supplementary Figure S9B. Estimated variable importance measures, i.e. % increase in mean squared error or MSE, from pre-grouped random forest implementation (RF #2) for the UK. Rows indicate cluster names (a full list of variables belonging to each cluster can be found in Supplementary Table S3) and corresponding principal components, if the cluster consists of multiple variables. PC1 denotes principal component 1 and PC2 denotes principal component 2 (sex-based population correction sensitivity analysis).**

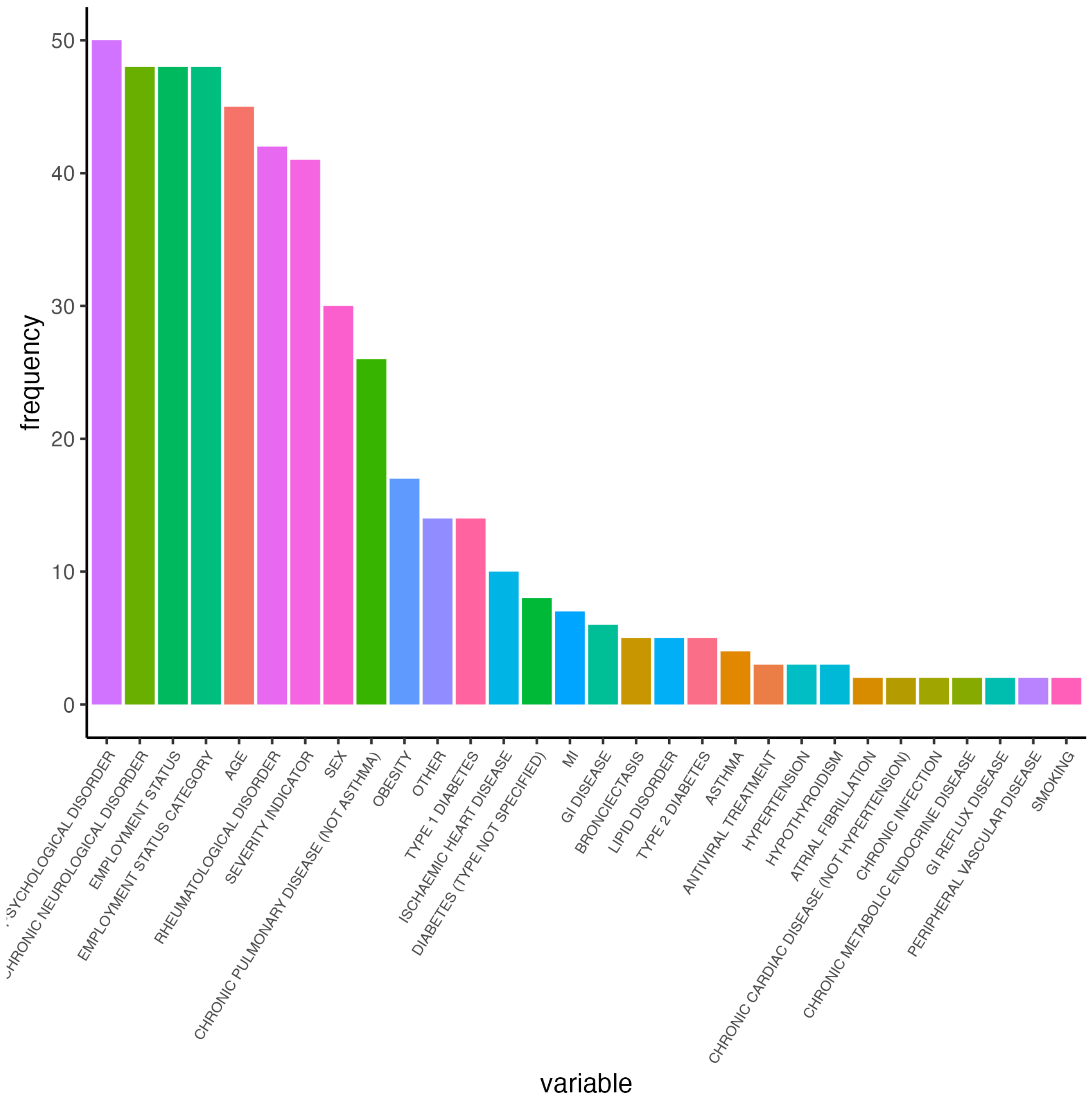


**Supplementary Figure S9C. Number of times (frequency) each variable appears in clusters selected for each CoV-VSURF run (RF #3) for the UK (sex-based population correction sensitivity analysis).**
